# Improving maternal outcomes of young mothers through mobile health (mHealth) and community-based interventions: Evidence from a quasi-experimental trial in Kwale County, Coastal Kenya

**DOI:** 10.1101/2025.10.27.25323180

**Authors:** Jefferson Mwaisaka, John Ganle, Adom Manu, Kwasi Torpey

## Abstract

**Background:** Inefficient referral systems and limited awareness on the importance of antenatal care (ANC) hinder expectant adolescents and young women from achieving the recommended ANC visits. Mobile health interventions and community-based clinical outreaches have emerged as potential strategies to improve ANC utilization and skilled delivery rates.

**Objective:** This study evaluates the effectiveness of mHealth text messaging and community-based clinical outreaches in improving ANC uptake, skilled deliveries, and infants’ birthweights among expectant adolescents and young women in Kwale County, Coastal Kenya.

**Methods:** A quasi-experimental study was conducted across four public health facilities, comparing three groups: mHealth text messaging intervention, community-based outreaches, and a control group receiving standard care. Multivariate regression models were used to adjust for baseline differences and potential confounders. Key outcomes included ANC attendance, skilled birth attendance, and infant birthweights.

**Results:** A total of 817 participants were included in the analysis. Participants in the mHealth (RR: 0.5 [95% CI: 0.4 – 0.7], p<0.001) and outreach arms (RR: 0.1 [95% CI: 0.0 – 0.7], p=0.024), had significantly lower risk of low ANC contacts compared to the control arm participants. Similarly, the odds of unskilled deliveries were significantly lower in the mHealth (OR: 0.2 [95% CI: 0.1 – 0.4, p<0.001] and the outreach arms (OR: 0.1 [95% CI: 0.0 – 0.6], p=0.007), compared to the control group. Notably, infants born to participants in the mHealth-text message intervention (OR: 5.9 [95% CI: 1.7 – 20.8, p=0.006]) and outreach arms (OR: 6.6 [95% CI: 1.7 – 25.7], p=0.007) had significantly higher odds of low birth weight, compared to those born to control arm participants

**Conclusions:** Both mHealth-text messaging and community-based clinical outreaches can improve antenatal contacts and institutional skilled deliveries. Integrating both strategies can enhance maternal health outcomes for young women in Kenya and should be considered for broader adoption within public health programs to optimize care delivery.

## Introduction

Each year, 2 million young girls aged 14 years and below and 21 million adolescent girls aged between 15-19 give birth in underdeveloped countries (1). Around 27% of young women in the least developed nations between the ages of 20 and 24 give birth before age 18 with Sub-Saharan Africa recording the greatest rate of early childbearing (2). In Kenya, an estimated 15% of girls aged 15-19 - approximately one in every seven are either pregnant or have given birth to their first child (3). The rising number of pregnant teenagers is a public health quandary because most of them avoid skillful antenatal health services hence resulting to maternal problems and unfavorable health outcomes (4). For instance, approximately 23.5% of adolescent mothers in Sub-Saharan Africa experience at least one adverse birth outcome, such as low birthweight, premature birth, or early neonatal mortality (5). These adverse outcomes are significantly associated with low-quality antenatal care (ANC). (5). The heightened risk of adverse birth outcomes among adolescent women is well-documented across various settings. In Kenya, the preterm birth rate among adolescent women was at one time reported at 5.6%, significantly higher than the 0.8% recorded among adult women (6). Inadequate pregnant women’s uptake of well-timed and regular prenatal care is a critical obstacle for improving prenatal indicators, and specifically among expectant young women (7). Additionally, there is enough scientific proof associating poor perinatal outcomes and other birth complications to inadequacy of ANC care among expectant adolescents and young women (4).

Evidence suggests that having at least four clinical antenatal contacts and consulting a competent healthcare provider during pregnancy is associated with a 3.09% lower risk of perinatal death and a 6.65% lower risk of delivering low-birth-weight infants. (8). A recent study in Sub-Saharan Africa reported that 50.8% of expectant adolescents received low-quality antenatal care (ANC), 32.4% received medium-quality care, and only 16.7% received high-quality care. Adolescents who received high-quality ANC had a 28% lower risk of experiencing adverse birth outcomes. (5). In Kenya, a study assessing the effectiveness of ANC opined that almost 40% of neonatal mortalities can be prevented by adherence to ANC appointments (9). Despite this association, the uptake of adequate and high-quality ANC remains low among adolescent girls and young women in Sub-Saharan Africa.

In Kenya, while 66% of all pregnant women complete at least four ANC contacts with a healthcare provider, only 57% of young mothers under the age of 20 receive the same level of care in 2022 (3). In Kwale County where this study was conducted, 72% of all pregnant women had at least four ANC contacts (3). However, an earlier study by Mwaniki et al., (2014) found that approximately two-thirds (67%) of expectant women, including adolescent girls and young women in Kwale County, do not adhere to their planned ANC schedules (10). Several factors have been attributed to low utilization of antenatal services in Kenya including poor and inadequate referral channels and poor quality of services provided to pregnant women (11). In Kenya, referral strategies have mainly relied on community health promoters to encourage positive antenatal health behavioural changes. This approach and the utilization of ANC appointment cards have been suboptimal in improving uptake of prenatal health services among adolescent girls and young women. Research in Kenya and Uganda found that securing the antenatal appointment card was seen by young women as a sure pass to receiving priority services when giving birth (12,13). For that reason, majority of expectant young girls and women in Kenya attend at least one prenatal clinic to ordinarily get the clinical appointment card. Also, a significant number of expectant young girls and women don’t know their clinical schedules even though the dates are usually written on their clinic booklets and cards; knowledge gaps among expectant young girls and women on the required number of clinical contacts have also been documented (12).

Following WHO’s recommendation to increase antenatal contacts to eight to reduce perinatal mortality and improve women’s experience of care (14), there is a need to design, test, and evaluate evidence-based interventions that effectively promote ANC service utilization among expectant young women, complementing existing demand-creation strategies. Community-based outreach programmes - an extension of primary healthcare services offered at health facilities and utilised to reach underserved communities have been found to have the potential to improve access to maternal services in Sub-Saharan Africa (15,16). Kenya also appreciates the power of community outreach programmes. The ministry of health in Kenya has developed an all-inclusive community health strategy outlining how coordinated outreaches and health awareness activities should be carried out (17). Additionally, the increased availability of cell phones and young women’s readiness to get health education messages via their mobile phones (18,19) point to the possibility that mHealth will change the way that formal healthcare is provided in low-income settings. mHealth interventions have shown potential to improve the quality, timeliness and coverage of health services (20–22). However, there is little evidence regarding the effectiveness of mHealth platforms targeting expectant young women aimed at improving ANC uptake and service utilization in sub-Saharan Africa including Kenya (23). Studies have also shown that SMS interventions have enhanced adherence to clinical appointments and hence contributing to timely seeking of healthcare services in several LMICs (24–28). However, few studies in East Africa, including Kenya, have reported similar findings on ANC uptake specifically among expectant young women and adolescent girls (23). Additionally, while mHealth (28–30) and community-based interventions (31,32) have been used to improve maternal outcomes, they often operate in silos and primarily target the general population rather than young mothers. This study was originally designed as a two-arm trial but was modified to three arms due to COVID-19 related declines in facility ANC attendance, which prompted the Kwale County Department of Health to introduce community outreach at one control facility. This unplanned introduction provided a unique opportunity to evaluate the effectiveness of both mHealth and community outreach interventions simultaneously, providing insights into how complementary interventions can improve maternal outcomes among young mothers in a real-world setting.

This quasi - experimental study therefore sought to determine the mHealth and community outreaches intervention’s effect on antenatal care utilization, institutional deliveries, and infants’ birthweights among expectant young women aged 24 and below in Kwale County, Coastal Kenya.

## Materials and Methods

### Study design

This was a quasi-experimental study with two arms to evaluate the impact of an mHealth text messaging intervention on ANC service utilization, institutional deliveries and infants’ birthweights among expectant young women (≤ 24 years) in Kwale County, Coastal Kenya. A quasi-experimental approach was chosen due to logistical constraints that precluded full randomization, including the need for geographical separation to minimize information spillover. The study was also conducted within the timeframe of a doctoral program, which required a design that balanced methodological rigor with feasibility. A randomized controlled trial would have required more time, extensive setup and resources beyond the scope of the PhD grant. This study was initially designed as a two-arm trial, with one arm receiving the intervention and the other serving as a comparison group. However, unintended deviations from the original study plan occurred due to the impact of the COVID-19 pandemic. The fear of contracting COVID-19 at health facilities led to a decline in ANC service uptake among expectant women. In response, the Kwale County Department of Health introduced community outreach activities to deliver critical maternal health services at the community level. As a result, one of the control facilities, Mkongani Health Centre, began conducting monthly community-based ANC outreaches. These outreaches provided maternal health education, screening, and ANC services, reaching 147 out of 213 study participants from Mkongani. Due to these external modifications, the original control arm was divided into two subgroups: (1) "pure controls," comprising participants from Mazeras Health Centre and Mkongani participants who did not attend outreaches, and (2) an "outreach arm," consisting of Mkongani participants who received ANC services through community outreach activities. Consequently, the study evolved into a three-arm design: (1) mHealth-text messaging intervention participants, (2) pure control participants, and (3) outreach participants. The study design was adapted accordingly to account for these modifications while maintaining methodological rigor. All subsequent analyses and results reflect this revised study design.

### Study setting

This study was conducted in Kwale County, Coastal Kenya, across four rural public health facilities which included Kikoneni, Mkongani, Mnyenzeni, and Mazeras health centres; selected from 11 eligible facilities based on the burden of teenage motherhood. Facilities with at least 200 young girls seeking pregnancy care in 2018 were identified as high-burden sites and served as a proxy for selecting study locations. A stratified random assignment was used to allocate health centres within this high-burden category (>200 teenage mothers in 2018) to either the intervention or control arm, ensuring that each facility had a single study arm. To enhance internal validity, facilities were selected to be comparable in terms of geographical access to healthcare, mobile network coverage, and operational capacity to implement the intervention.

### Study Population and Eligibility Criteria

The study population comprised expectant young women aged 24 years and below attending their first ANC clinic at one of the four selected public health facilities. Eligible participants were required to reside within the study area with no travel plans during the study period, have a gestational age of 24 weeks and below, own a mobile phone at the time of recruitment, and possess basic literacy in English or Swahili. The gestational age cut-off was based on a nationally representative survey in Kenya, which reported a mean gestational age of 23.8 weeks at first ANC contact among expectant young women (33). Participants with an uneventful first trimester were enrolled after providing informed consent. Expectant young women presenting with pregnancy-related complications at their first ANC visit, including hyperemesis gravidarum, placenta previa, gestational hypertension, gestational diabetes mellitus, pre-eclampsia, or iron deficiency anemia were excluded from the study. Frequency matching was applied over individual matching to ensure balance between groups on key characteristics while maintaining flexibility in recruitment. This approach allowed us to minimize confounding by age, gestational age, health centre accessibility, and network availability, without the logistical constraints of one-to-one matching.

### Intervention description

Messages from Mission Motherhood – (A non-profit organization offering mental and digital support to expectant and new mothers to safeguard their health and their babies’ well-being in the first thousand days of life) - (34) were used in this study. Messages were translated from English to Swahili to enhance comprehension and relate to the targeted participants. Prior to implementation, Focus Group Discussions with expectant adolescents were conducted to refine and validate the messages for clarity and relatability. The intervention primarily consisted of personalized, gestational age-specific SMS messages delivered via an mHealth platform. Upon enrolment, all participants in the intervention group received a welcome message regardless of their gestational age. Educational messages tailored to their specific gestational age were sent throughout the pregnancy, with frequency increasing as pregnancy progressed. In line with WHO’s ANC recommendations of a minimum of eight ANC contacts, one in the first trimester, two in the second trimester, and five in the third trimester; participants received one educational message per month during the first trimester, two messages per month in the second trimester, and weekly messages from week 27 until childbirth. Automated reminder messages were also sent 24 hours before scheduled ANC visits. The intervention was implemented at Kikoneni and Mnyenzeni Health Centres (mHealth intervention group, n = 398, 48.7%), while Mazeras and Mkongani Health Centres served as the control group (n = 419, 51.3%), receiving standard ANC services without SMS support. All messages were delivered in Swahili, as preferred by all participants. To address cultural barriers and stigma associated with low ANC uptake, participants had the option to register a spouse or caregiver as a "treatment buddy," who received supplementary monthly educational messages. This component was voluntary and not a criterion for study inclusion. Figs 1 and 2 shows study design flow diagram and the intervention logic model respectively.

**Figure 1.**
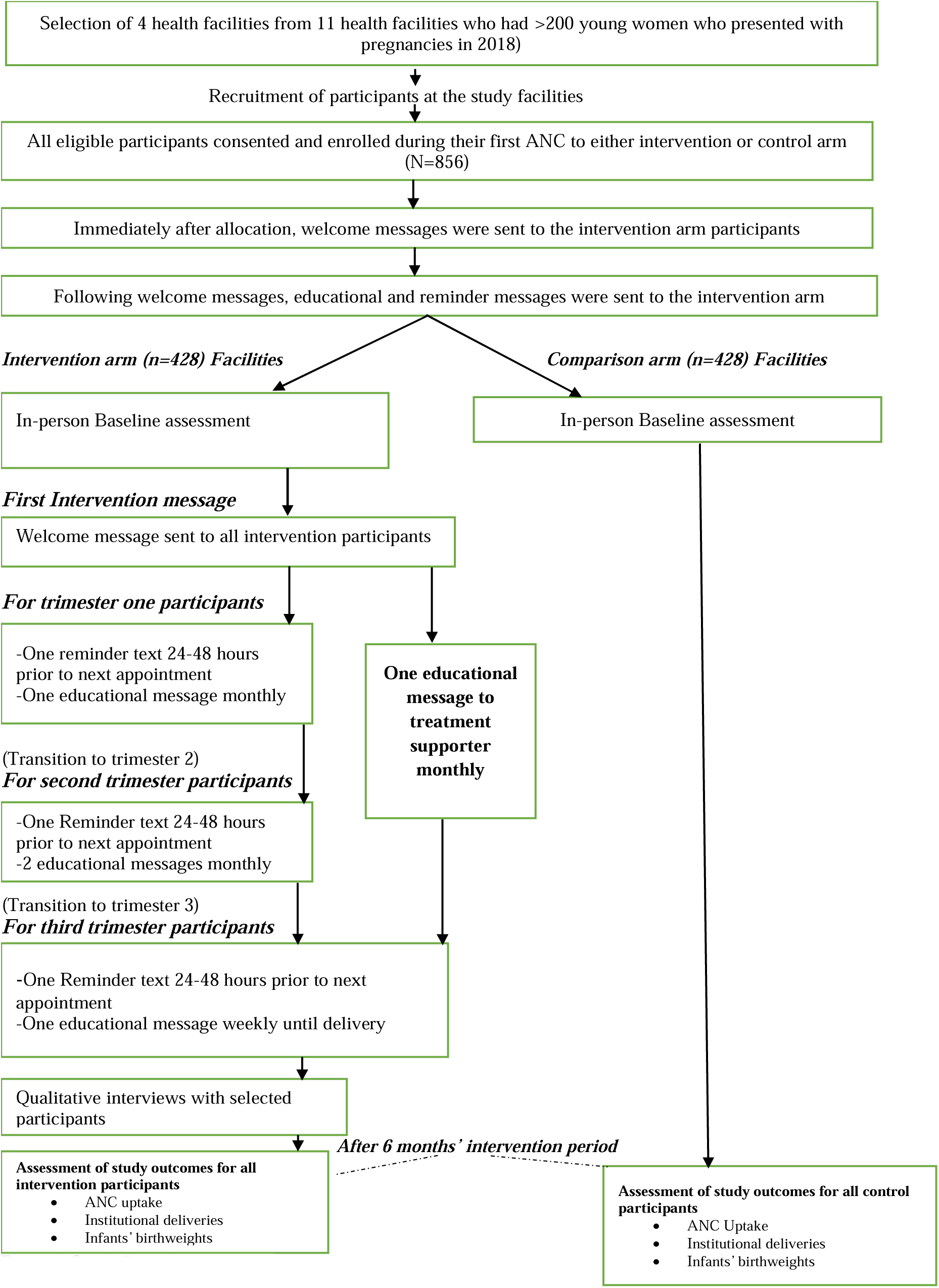
Study flow diagram: The diagram shows the flow of participants through each stage of the study, including recruitment, enrollment, allocation, text messaging by trimester for mHealth intervention participants, and assessment of study outcomes.

**Figure 2.**
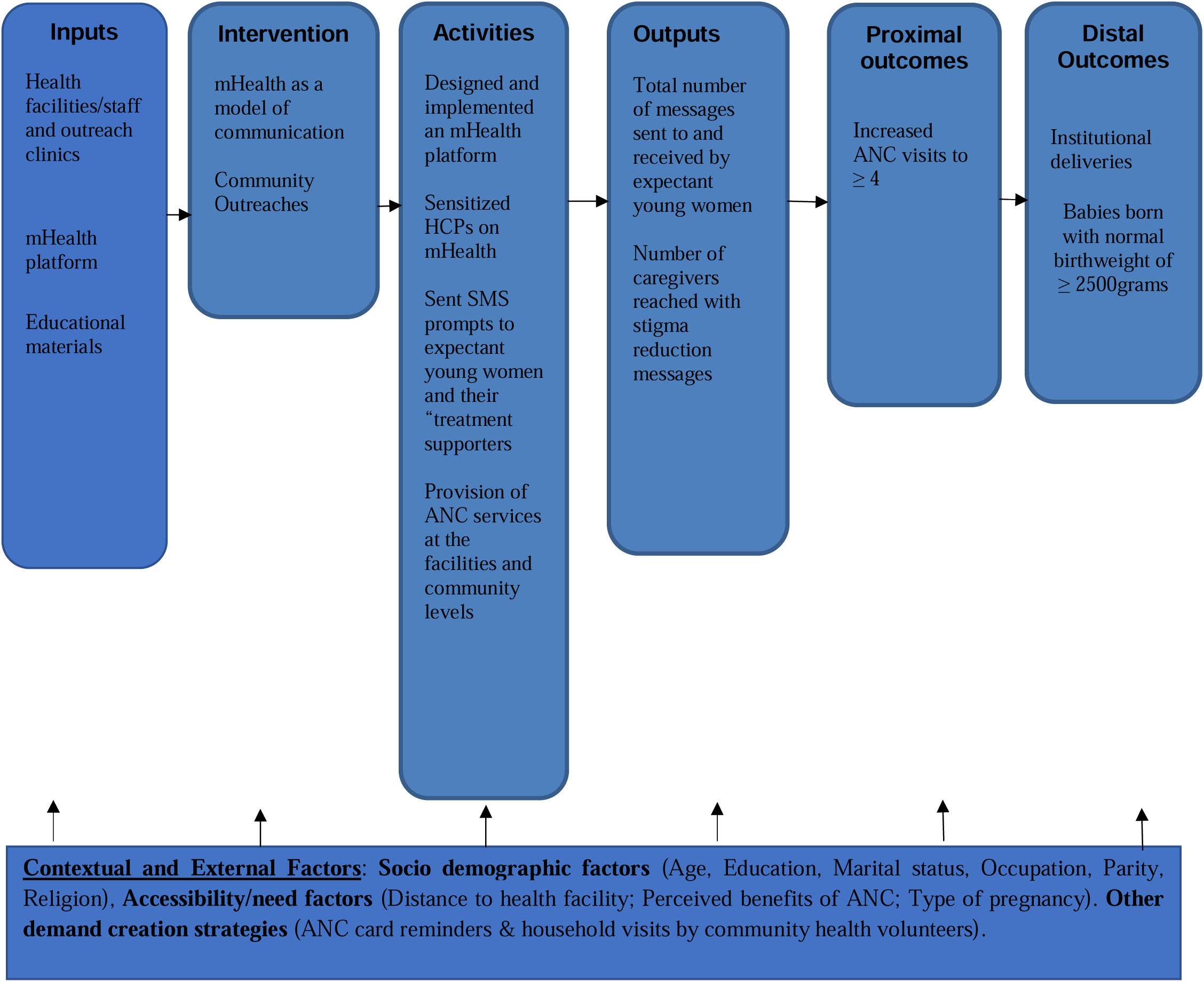
Intervention logic model: This model illustrates the pathways through which the mHealth and community outreaches’ intervention components are expected to influence the proximal and distal outcomes.

### Sample size and participants recruitment

The sample size estimation was based on the proportion of expectant teenagers in Kenya who did not attend at least four ANC visits, which was estimated at 51% (35) at the time this study was being conceptualized. To assess the impact of the intervention, a 10% increase in the proportion of expectant young women receiving at least four ANC contacts, as recommended by Kenya’s Reproductive, Maternal, Newborn, Child, and Adolescent Health (RMNCAH) guidelines, was considered clinically significant. Assuming this 51% represented the intervention group receiving text messages, a sample size of 778 participants (389 per arm) was calculated to achieve 80% power to detect a 10% difference at a 95% confidence level (α = 0.05). To account for an estimated 10% loss to follow-up, the study sample size was set at 856 expectant young women, with 428 participants allocated to each of the two initial study arms (mHealth and comparison).

Participants were recruited at health facilities during their first ANC visit within the study period. Healthcare providers (HCPs) informed expectant young women about the study after they received routine ANC services. As the first point of contact, HCPs identified eligible participants and introduced them to the study. Those who consented to participate were then linked to trained female research assistants stationed at the study facilities, who facilitated the written informed consent process and data collection. The ethical committee granted a waiver for minors who were pregnant, as they are considered emancipated minors in Kenya. The study was conducted from 16^th^ September 2020 to 18^th^ July 2021.

### Data collection procedures

Participants’ background characteristics data was collected using an interviewer-administered questionnaire, capturing demographic details, pregnancy-related information and ANC knowledge. Data was collected electronically using Open Data Kit (ODK) by trained female data collectors who conducted face-to-face interviews while adhering to COVID-19 prevention measures. Post-trial data was extracted from the Ministry of Health’s ANC registers (MOH 405) and maternity registers (MOH 333). The ANC registers provided information on the number of ANC visits, gravidity, gestational age, estimated delivery date, and any pregnancy complications detected or treated. The maternity registers contained data on delivery outcomes, including gestational age at birth, mode of delivery, maternal condition post-delivery, and neonatal details such as birth weight, birth outcomes, deformities, and the attending healthcare provider. Referral records were also documented. Participants who did not deliver at study facilities were contacted via phone for follow-up. Unique ANC numbers were recorded throughout the study to link pre-intervention and outcome data for analysis.

### Data analysis

The primary outcome of the study was an increase in antenatal contacts to four or more, defined as the total number of clinical ANC visits made by study participants from enrolment until delivery, categorized as 0 for less than 4, and 1 for four or more ANC contacts. Secondary outcomes included institutional deliveries, defined as childbirths occurring in a health facility under the care of a skilled birth attendant, measured as 0 for unskilled deliveries, and 1for institutional deliveries. Infant birthweight was also a secondary outcome, defined as the baby’s weight immediately after birth, categorized as low birthweight (<2,500 grams) or normal birthweight (≥2,500 grams).

Descriptive statistics were used to summarize participants’ background characteristics, with means and standard deviations presented for continuous variables and proportions for categorical variables. Categorical variables were checked for differences between the three groups using chi square tests. The Kruskal-Wallis test was used to determine if there was a statistically significant mean difference between the three groups regarding antenatal contacts, institutional deliveries and infant birth weights. Analysis of Covariance was done to control for any unanticipated pre-existing differences between the three study groups based on their pre-intervention background characteristics. This was followed by multivariate logistic regression analysis on the study binary outcomes of (1) four or more antenatal contacts (yes or no), (2) Institutional deliveries (yes or no), and (3) infant birthweights (low birthweight =<2500GRAMS or normal birthweight infants >2500grams) to assess the effect of the mHealth-text messaging and outreach interventions. Multivariate logistic regression with a 95% confidence interval was used to assess the relationships between independent variables and the study’s outcomes. Adjusted analyses controlled for age, education, religion, age at sexual debut, marital status, pregnancy planning, parity, and time to facility, based on observed differences across intervention arms in this study. No formal statistical matching was conducted during analysis; all participants were included in the analyses. Frequency matching was applied only at the design stage to ensure comparable groups on age, gestational age, health centre accessibility, and network availability. Findings are presented as odds and risk ratios. Results were statistically significant at p<0.05. All quantitative data analyses were conducted using STATA version 15 (StataCorp. 2017. *Stata Statistical Software: Release 15*. College Station, TX: StataCorp LLC).

## Ethical considerations

This study was approved by the Noguchi Memorial Institute for Medical Research Institutional Review Board at the University of Ghana, (NMIMR-IRB CPN 066/19-20), the University of Nairobi, Ethics and Review Committee at the Kenyatta National Hospital (KNH-UoN ERC) KNH-ERC/A/234), and the National Commission for Science, Technology and Innovation (NACOSTI) (NACOSTI/P20/6389). Permission to conduct the research was sought from the Kwale County Health Management Team and in-charges of the four health facilities involved in the study. All participants gave informed consent to participate.

## Results

The mean age at first ANC visit was 21.1 years [standard deviation-SD, 2.4]), at sex debut 17.4 [SD, 2.2] and at first pregnancy 18.8 years [SD, 2.5]). The mean age of the spouse/partner was 26.8 years (SD, 4.8). Majority of participants had primary level education (82.2%), were Muslim (71.1%), had their sexual debut at more than 18 years (51.3%), and were engaged or married (83.7%). Additionally, majority of the participants in this study were neither formally employed nor doing business (90.8%), had a male partner as their primary caregiver (79.2%), reported that their pregnancy was planned (72.0%), said that this was their first pregnancy (57.2%) and also reported that their age at first pregnancy was above 20 years (61.0%). Only 12.1% of all participants attended their first ANC visit in the first trimester. The majority were over 20 weeks pregnant at their first ANC visit (70.0%), reported to have received some ANC information since becoming pregnant (60.1%), and reported a travel time of more than 30 minutes to reach their respective health facilities. (70.5%). Table 1 below presents the primary characteristics relevant to the analysis. The full dataset is available in Supplementary Table 1.

**Table 1:**
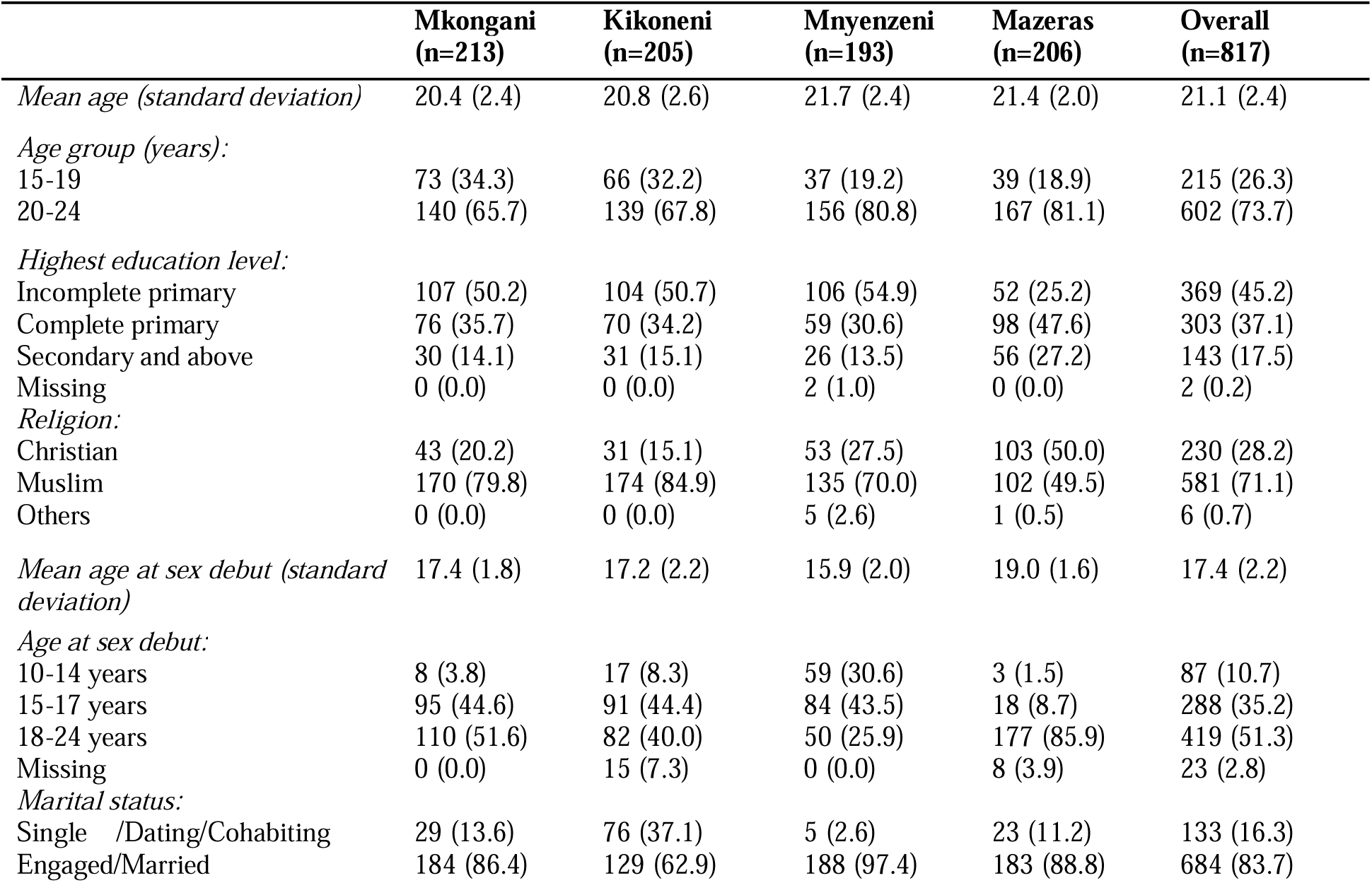
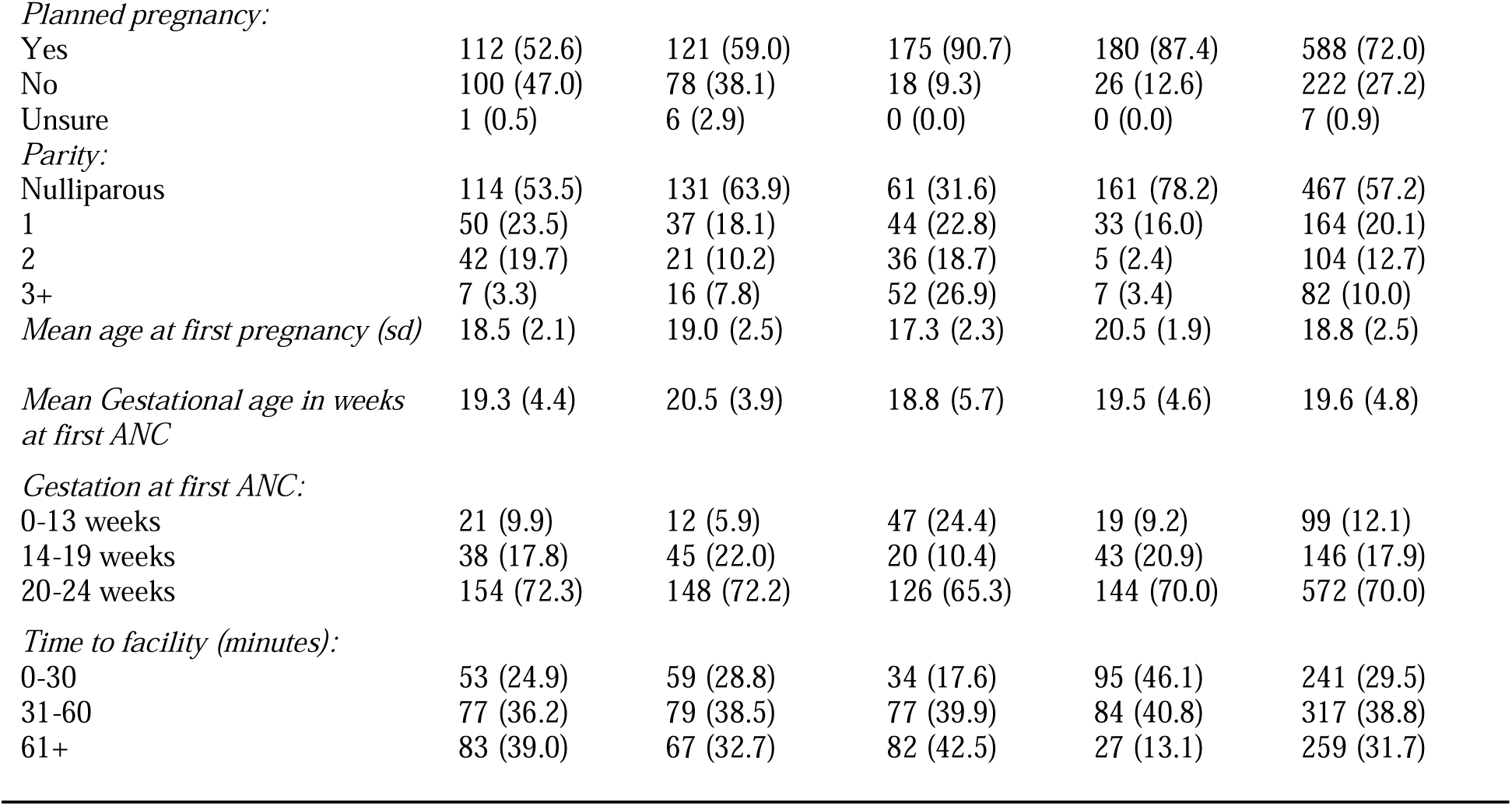
Background characteristics of study participants (N=817)

Compared to participants from the control and outreach arms, participants from the mHealth intervention arm were more likely to have an incomplete primary school education (52.8% vs control 31.6% and outreach 49.7%, p<0.001), to have a younger age at sexual debut (less than 15 years, 19.1% vs control 2.2% and outreach 3.4%, p<0.001) and to be single/dating/cohabiting (20.4% vs. control 10.7% and outreach 15.7%, p=0.002). mHealth intervention participants were also more likely to be supported by their parents (25.6% vs control 10.3% and outreach 13.6%, p<0.001), to be employed or doing business (11.6% vs. control 8.1% and outreach 4.8%, p=0.04) and to have the current pregnancy as their third or more pregnancy (17.1% vs control 3.7% and outreach 2.7%, p<0.001). Moreover, mHealth intervention participants were more likely to have been less than 18 years at their first pregnancy (40.5% vs control 11.8% and outreach 26.5%, p<0.001) and to have received any ANC information since becoming pregnant (80.4% vs control 46.0% and outreach 31.3%, p<0.001). Further, the outreach arm participants were more likely to be Muslims (78.2% vs. control 57.7% and intervention 77.6%, p<0.001), and more likely to take longer time to get to the facility from home (more than 60 minutes, 42.2% vs. control 17.7% and intervention 37.4, p<0.001). Control participants were more likely to be having a planned pregnancy (80.9% vs intervention 74.4% and outreach arm 49.0%, p<0.001). Table 2 below presents a comparison distribution of study participants by their intervention status. The full comparison dataset is available in Supplementary Table 2.

**Table 2:** A comparison distribution of study participants by their intervention status (N=817)

| Characteristics | Control<br>(n=272) | Intervention<br>(n=398) | Outreach<br>(n=147) | p-value* |
| --- | --- | --- | --- | --- |
| *Median age (IQR)* | 21 (20-23) | 21(19-24) | 20 (19-22) | <0.001 |
| *Age group (years):* |  |  |  |  |
| 15-19 | 55 (20.2) | 103 (25.9) | 57 (38.8) |  |
| 20-24 | 217 (79.8) | 295 (74.1) | 90 (61.2) | <0.001 |
| *Highest education level:* |  |  |  |  |
| Incomplete primary | 86 (31.6) | 210 (52.8) | 73 (49.7) |  |
| Complete primary | 118 (43.4) | 129 (32.4) | 56 (38.1) |  |
| Secondary and above | 68 (25.0) | 57 (14.3) | 18 (12.2) |  |
| Missing | 0.0 | 2 (0.5) | 0.0 | <0.001 |
| *Religion:* |  |  |  |  |
| Christian | 114 (41.9) | 84 (21.1) | 32 (21.8) |  |
| Muslim | 157 (57.7) | 309 (77.6) | 115 (78.2) |  |
| Others | 1 (0.37) | 5 (1.3) | 0.0 | <0.001 |
| *Median age at sex debut (IQR)* | 19 (18-20) | 16 (15-18) | 18 (16-18) | 0.0001 |
| *Age group at sexual debut:* |  |  |  |  |
| 10-14 years | 6 (2.2) | 76 (19.1) | 5 (3.4) |  |
| 15-17 years | 48 (17.7) | 175 (44.0) | 65 (44.2) |  |
| 18-24 years | 210 (77.2) | 132 (33.2) | 77 (52.4) |  |
| Missing | 8 (2.9) | 15 (3.8) | 0.0 | <0.001 |
| *Marital status:* |  |  |  |  |
| Single/Dating/Cohabiting | 29 (10.7) | 81 (20.4) | 23 (15.7) |  |
| Engaged/Married | 243 (89.3) | 317 (79.7) | 124 (84.4) | 0.004 |
| *Planned pregnancy:* |  |  |  |  |
| Yes | 220 (80.9) | 296 (74.4) | 72 (49.0) |  |
| No | 51 (18.8) | 96 (24.1) | 75 (51.0) |  |
| Unsure | 1 (0.4) | 6 (1.5) | 0.0 | <0.001 |
| *Parity:* |  |  |  |  |
| Nulliparous | 195 (71.7) | 192 (48.2) | 80 (54.4) |  |
| 1 | 47 (17.3) | 81 (20.4) | 36 (24.5) |  |
| 2 | 20 (7.4) | 57 (14.3) | 27 (18.4) |  |
| 3+ | 10 (3.7) | 68 (17.1) | 4 (2.7) | <0.001 |
| *Median age at first pregnancy (IQR)* | 20 (18-21) | 18 (16-20) | 18 (17-20) | 0.0001 |
| *Gestation at first ANC:* |  |  |  |  |
| 0-13 weeks | 26 (9.6) | 59 (14.8) | 14 (9.5) |  |
| 14-19 weeks | 56 (20.6) | 65 (16.3) | 25 (17.0) |  |
| 20-24 weeks | 190 (69.9) | 274 (68.8) | 108 (73.5) | 0.151 |
| *Time to facility (mins):* |  |  |  |  |
| 0-30 | 115 (42.3) | 93 (23.4) | 33 (22.5) |  |
| 31-60 | 109 (40.1) | 156 (39.2) | 52 (35.4) |  |
| 61+ | 48 (17.7) | 149 (37.4) | 62 (42.2) | <0.001 |
| *Continuous variables assessed using Kruskal-Wallis test; categorical variables assessed using the chi-square test |  |  |  |  |

For all outcomes, adjusted analyses controlled for age, education, religion, age at sexual debut, marital status, pregnancy planning, parity, and time to facility, based observed differences across intervention arms and on prior evidence of their influence on maternal health outcomes, as shown in table 2.

### Effect of text message and outreach interventions on ANC contacts

Participants made a median of 4 (Interquartile range (IQR), 3 - 5) ANC contacts. Overall, 273 (33.4% [95% CI: 30.2% – 36.8%]) participants had low (**<** 4) ANC contacts. Among the three study arms, participants in the outreach arm had a significantly lower prevalence of low ANC contacts (2% [95% CI: 1%-6%]) compared to mHealth text message intervention arm (30% [95% CI: 25%-35%]) and control arm which had the highest prevalence of low ANC contacts (55% [95% CI: 49% – 61%]), p<0.001).

In a multivariate regression model and after controlling for covariates, participants in the mHealth text-message intervention arm (Risk Ratio, RR: 0.5 [95% CI: 0.4 – 0.7], p<0.001) and outreach arm (RR: 0.1 [95% CI: 0.0 – 0.7] p = 0.024, had a significantly lower risk of low ANC contacts compared to participants in the control arm (Table 3). Other factors that were associated with low ANC contacts included whether the pregnancy was planned and gestation at first ANC contact. Specifically, participants with a planned pregnancy had significantly higher risk (higher likelihood) of low ANC contacts compared to those with an unplanned pregnancy (RR, 2.3 [95% CI: 1.5 – 3.5], p<0.001). There was also an increase in the prevalence of low ANC contacts with increasing gestation at first ANC visit. Specifically, participants attending their first ANC clinic at 20 or more weeks of gestation had significantly increased risk of low ANC contacts compared to those attending their first ANC clinic at 13 or less weeks of gestation (RR, 1.6 [95% CI: 1.2 – 2.1], p=0.003). Table 3 presents effect of mHealth text message and outreach interventions on ANC contacts. The full analysis is available in Supplementary Table 3.

**Table 3:**
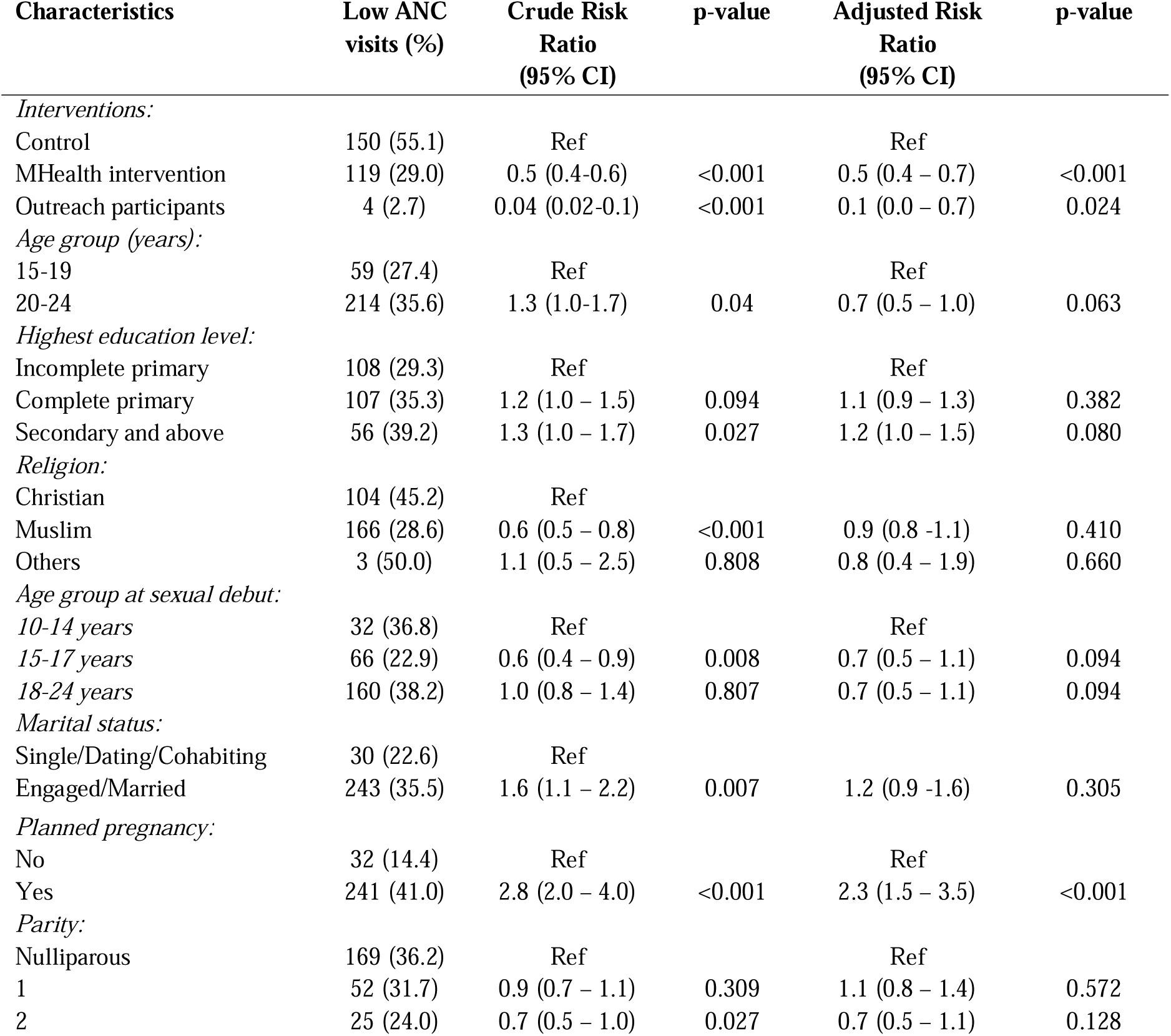
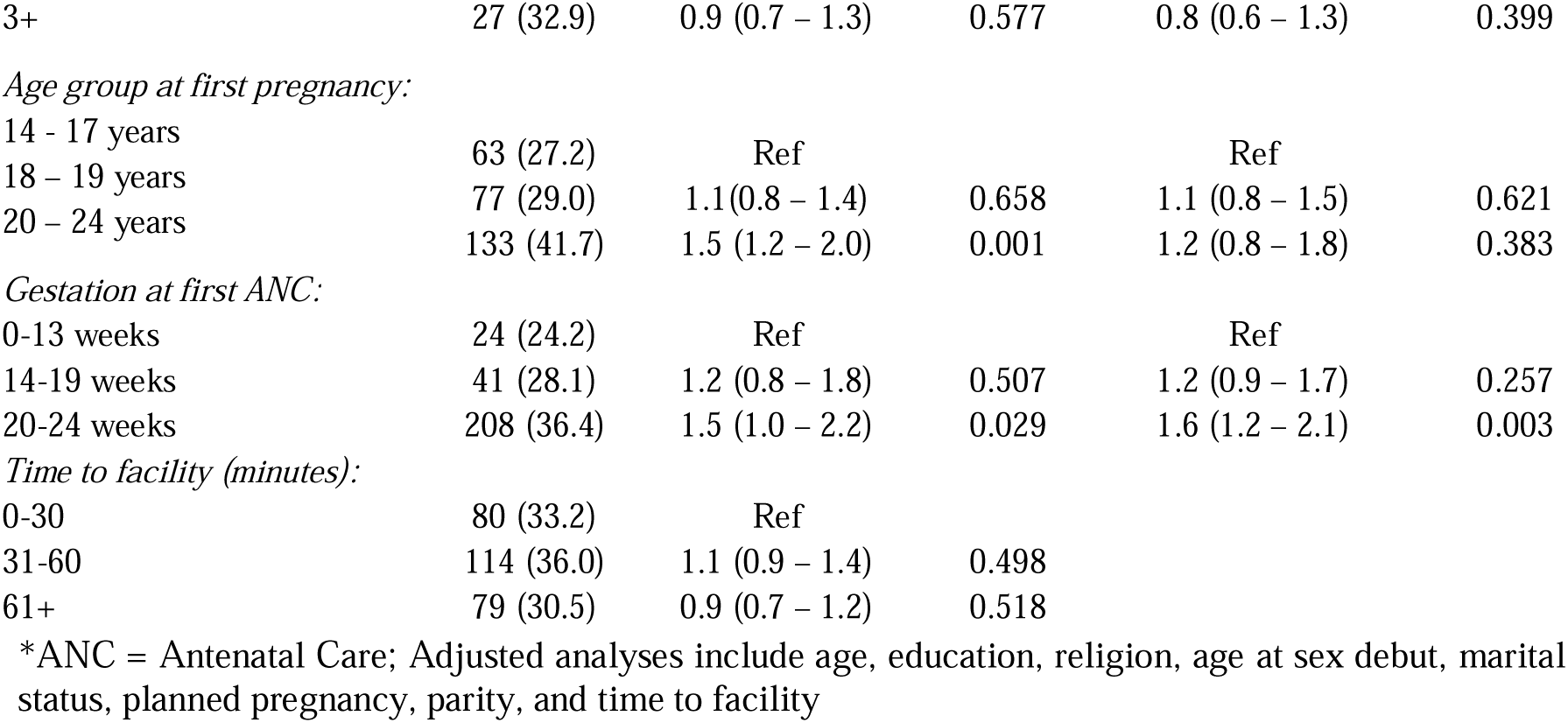
Effect of mHealth text message and outreach interventions on ANC contacts amongst study participants (N=817)

### Effect of text message and outreach interventions on skilled deliveries

Of the 817 participants included in the analysis, 25 (3.1%) were missing data on birth attendant and were excluded from this sub-analysis. Of the remaining 792 participants, 31 (3.9% [95% CI: 2.6% – 5.5%]) were delivered by unskilled traditional birth attendants. Compared to participants from the control arm, those from the mHealth-text message intervention and outreach arms had significantly lower prevalence of unskilled deliveries (9.0% [95% CI: 6.0% – 13.4%] vs. mHealth-text message intervention arm 1.2% [95% CI: ∼1.0% – 3.0%] and outreach arm 1.4% [95% CI: ∼1.0% – 4.9%], p<0.001).

In a multivariate regression model and after controlling for covariates, participants in the mHealth-text message intervention arm (OR: 0.2 [95% CI: 0.1 – 0.4, p<0.001] and the outreach arm (OR: 0.1 [95% CI: 0.0 – 0.6], p=0.007) had significantly lower odds of unskilled deliveries compared to participants in the control arm. The only other factor that was associated with unskilled deliveries was the level of education. Specifically, participants who had completed primary education as their highest level of education had significantly higher odds of unskilled deliveries compared to those who had incomplete primary education (OR, 3.2 [95% CI: 1.2 – 8.2], p=0.019), though these results should be interpreted with caution because of the small numbers in both categories. Table 4 presents effect of mHealth text message and outreach interventions on skilled deliveries. The full analysis is available in Supplementary Table 4.

**Table 4:**
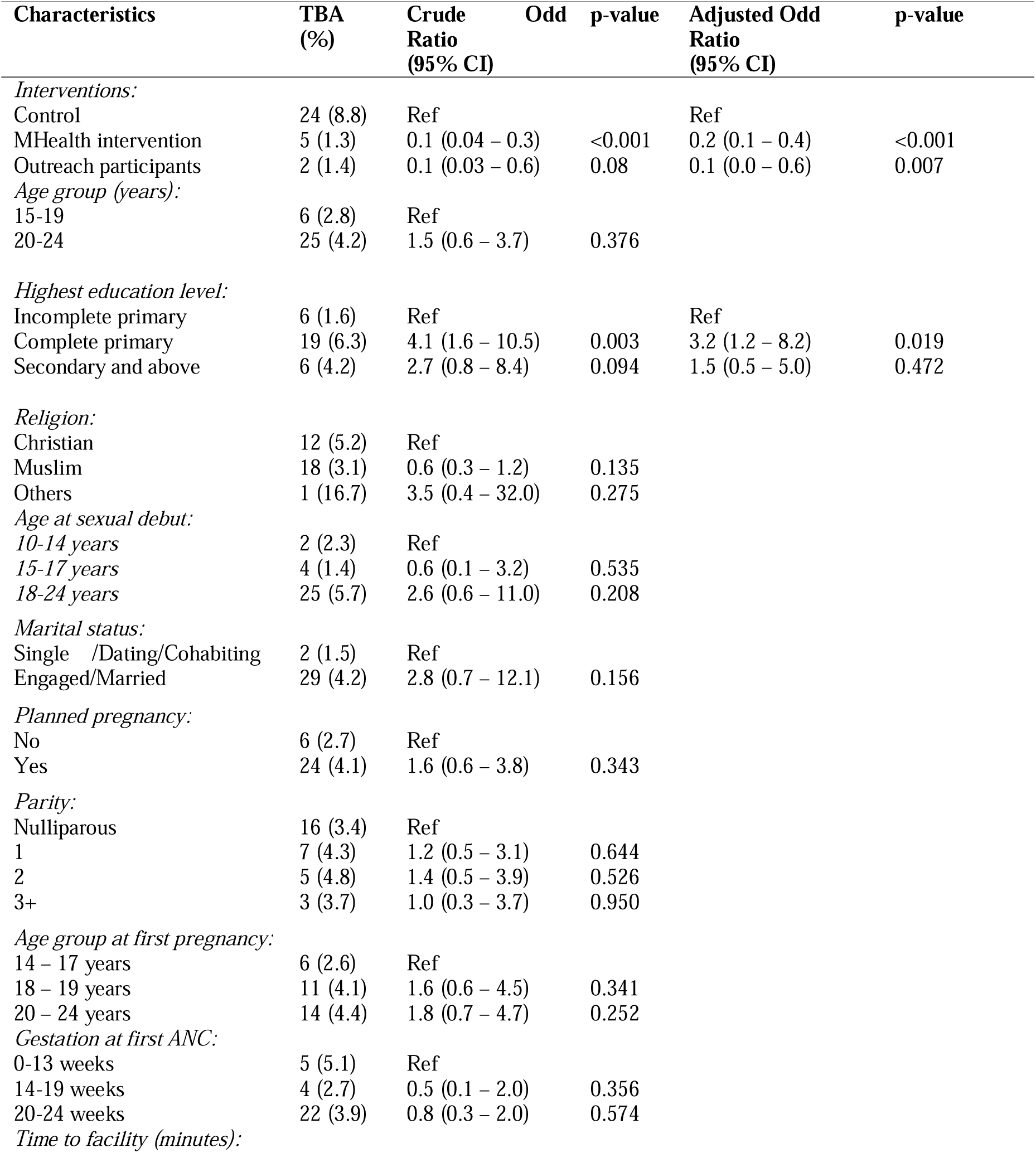
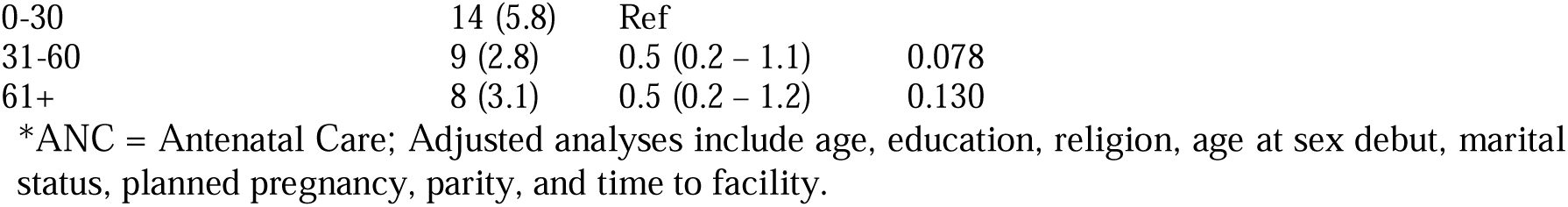
Effect of text messaging and outreach interventions on skilled deliveries

### Effect of text message and outreach intervention on infants’ birth weight

Of the 817 participants included in the analysis, 28 (3.4%) had babies who were missing data on birth weight and were excluded from this analysis. Of the remaining 789 participants, 51 (6.5% [95% CI: 4.9% – 8.4%]) had babies with a low (<2,500 grams) birth weight while 738 (93.5%) had normal birthweights. Compared to babies born to participants from the control arm, those born to participants from the mHealth-text message intervention and outreach arms had a significantly higher prevalence of low birth weights (1.1% [95% CI: ∼1.0% – 3.3%]) vs. mHealth-text message intervention arm 10.1% [95% CI: 7.3% – 13.6%]) and outreach arm (6.2% [95% CI: 2.9% – 11.5%]), p<0.001).

In a multivariate regression model and after controlling for covariates, infants born to participants from the mHealth-text message intervention (OR: 5.9 [95% CI: 1.7 – 20.8, p=0.006]) and outreach arms (OR: 6.6 [95% CI: 1.7 – 25.7], p=0.007) had significantly higher odds of low birth weight, compared to those born to control arm participants. The other factors that were associated with low birth weight were parity, whether participants received any ANC information since becoming pregnant and participants’ knowledge on when to start their ANC clinic. Specifically, participants who reported a parity of three or more had significantly lower odds of delivering low birth-weight babies compared to nulliparous participants (OR: 0.2 [95% CI: 0.0 – 0.8], p=0.026]). Participants who received some ANC information after getting pregnant had significantly higher odds of delivering low birth weight babies compared to those who did not receive any ANC information after getting pregnant (OR, 3.3 [95% CI: 1.4 – 8.2], p=0.008). Participants’ knowledge of when to start their ANC clinic was also significantly associated with low birthweight babies. Compared to participants who reported that pregnant women should start their antenatal clinics on missing their monthly periods. Those who responded that they should start at the first trimester ((OR, 0.3 [95% CI: 0.1 – 0.7], p=0.004), second trimester (OR, 0.3 [95% CI: 0.1 – 0.8], p=0.021) and those who didn’t know (OR, 0.2 [95% CI: 0.0 – 0.9], p=0.034) had significantly lower odds of delivering low birthweight babies. These results should also be interpreted with caution because of the small numbers in both categories. Table 5 presents effect of mHealth text message and outreach interventions on infants’ birthweights. The full analysis is available in Supplementary Table 5.

**Table 5:**
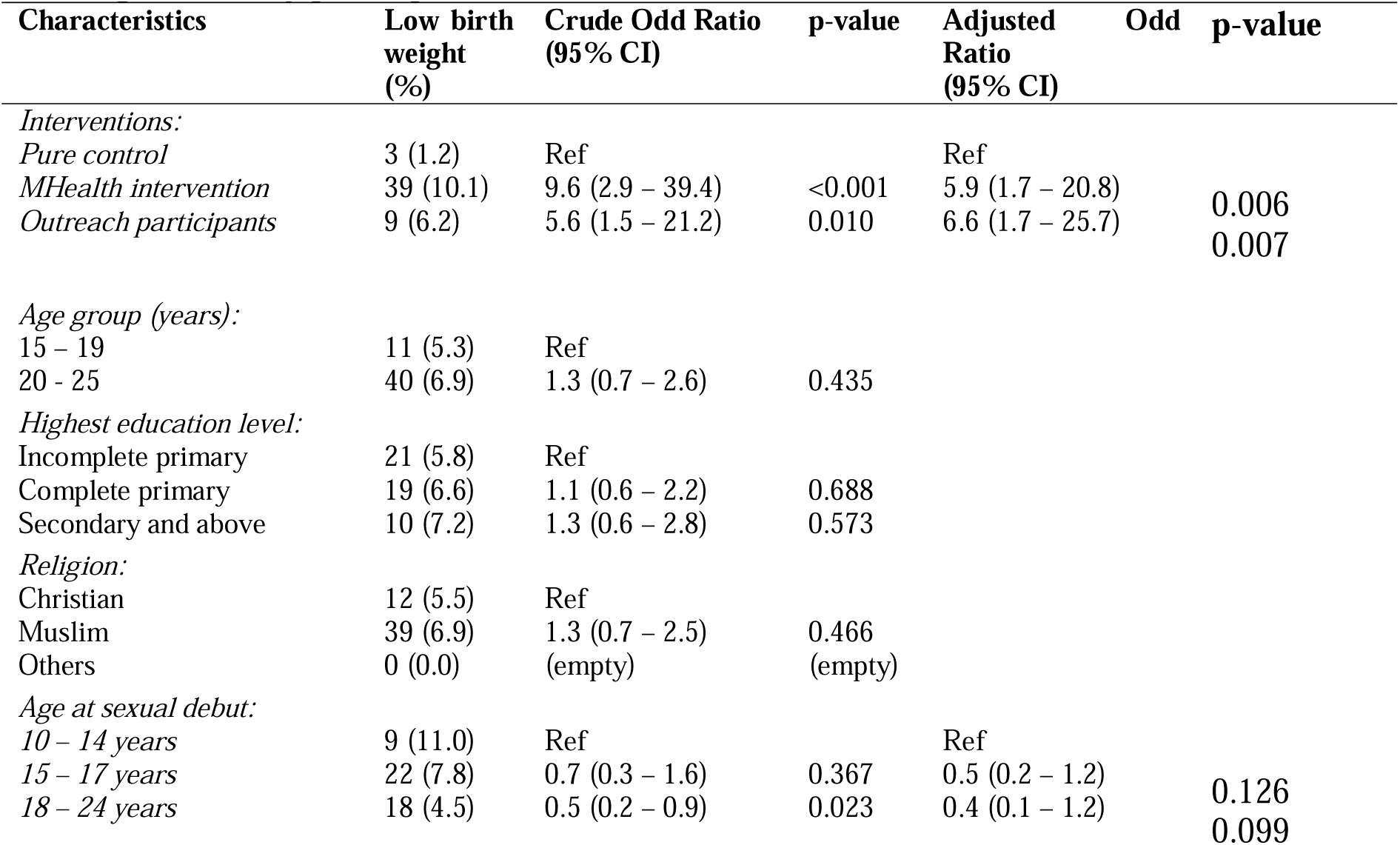
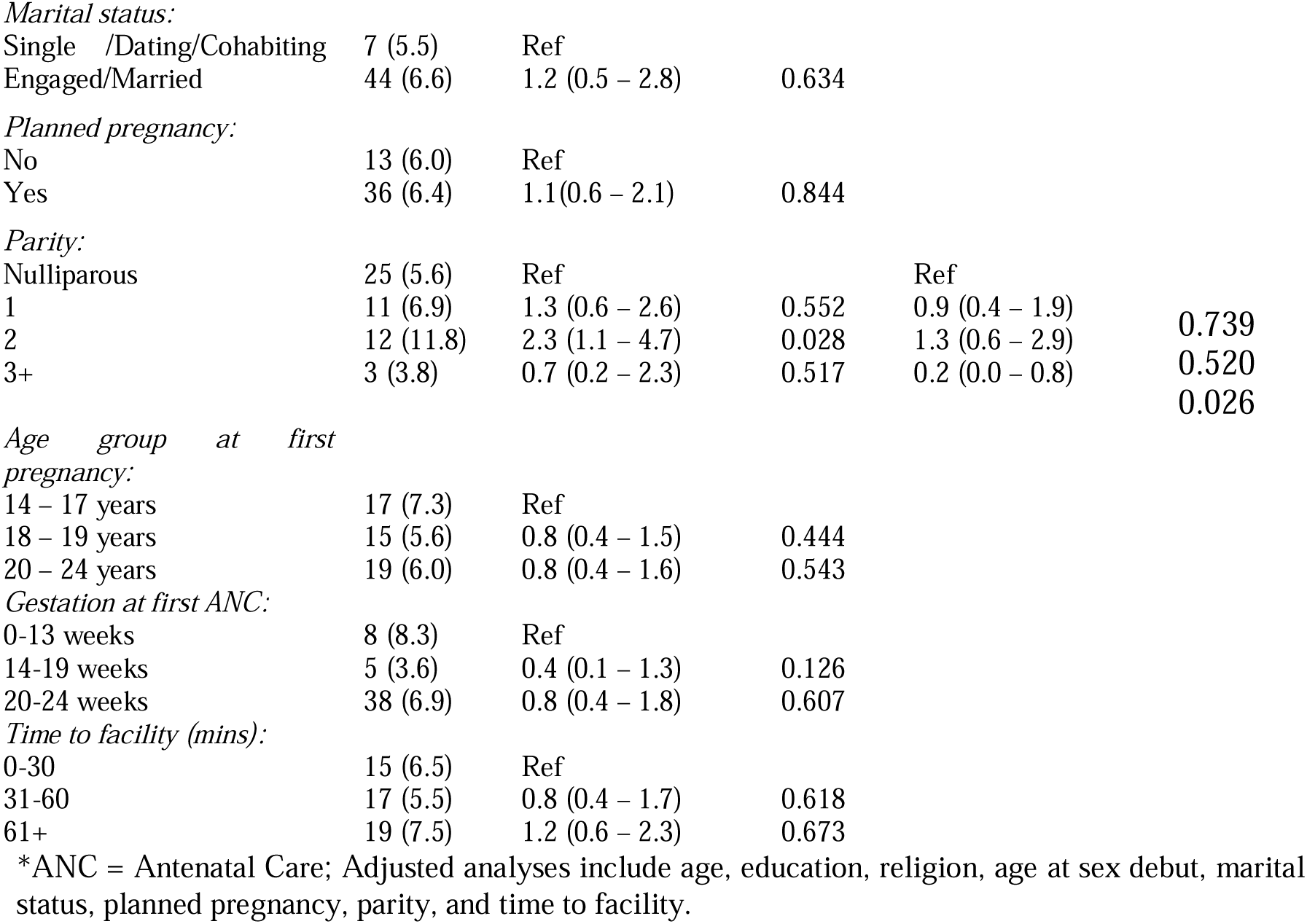
Effect of text messaging and outreach interventions on birthweight (<2500) amongst the study participants

## Discussion

This study found that only 12.1% of participants attended their first ANC visit in the first trimester, and 33.4% had fewer than four ANC contacts, falling short of Kenya’s Ministry of Health guidelines (36). These findings highlight delayed and suboptimal ANC utilization among expectant young women in Kwale County. They are consistent with prior research findings in Kenya and Sub-Saharan Africa (4,37–39) underscoring a unique vulnerability: adolescents and young women remain at heightened risk of inadequate maternal care despite policies promoting early and frequent ANC. Late ANC initiation (≥20 weeks) was significantly associated with fewer ANC contacts, supporting existing evidence that early ANC uptake is critical for achieving adequate antenatal care (39,40).

Interestingly, while previous studies identified education as a key determinant of early ANC uptake (4), this study did not find a significant association between secondary education and early ANC uptake. This may be due to the relatively low educational attainment across the sample (∼82% had only primary or no formal education), but it also raises questions about whether structural and sociocultural barriers, such as stigma, gendered decision-making, or lack of autonomy, may override the protective effect of education in this context. Moreover, consistent with other studies (4,38,39,41), limited knowledge about importance antenatal care continues to act as a barrier, underscoring the need for community-wide sensitization on the benefits of early ANC uptake, particularly for expectant adolescents, who face heightened risks of pregnancy-related complications.

A particularly unexpected finding was the significant association between planned pregnancy and low antenatal contacts. Previous evidence typically links unplanned pregnancies with inadequate antenatal uptake (4,40), but this finding aligns with evidence from Sub-Saharan Africa showing inconsistent patterns (42). One interpretation is that women with planned pregnancies may perceive themselves as being at lower risk, delaying care-seeking as a result. Alternatively, sociocultural influences, such as prevailing community norms regarding the appropriate timing for initiating care, may exert a stronger influence on health-seeking behavior than pregnancy intention alone (43). This contradiction calls for further qualitative work to unpack how perceptions of risk, pregnancy planning, and community expectations interact to influence ANC behavior among expectant adolescents and young women.

This study also demonstrated that both mHealth and outreach interventions improved antenatal contacts. This finding reinforces evidence that SMS-based interventions can positively influence care-seeking by providing reminders, education, and encouragement (20,24,44–46). However, behavioral change among adolescents is complex, often constrained by stigma, social norms, or household-level decision-making. As such, mHealth alone may be insufficient, requiring complementary approaches to address broader barriers. The unplanned introduction of outreach, prompted by COVID-19 disruptions, offered a natural experiment that added depth to the study. While this study was not originally designed to assess the effectiveness of community outreaches in improving antenatal uptake, the analysis indicates that such interventions hold promise for increasing antenatal uptake among expectant young women. Outreach participants were 90% less likely to have fewer than four antenatal contacts compared to controls, consistent with findings from other Sub-Saharan African settings where community-based clinical outreaches improved maternal health knowledge and service use (31,47,48). The presence of healthcare providers in community level reduced access barriers and built trust with young mothers. This suggests that integrating community-based clinical outreaches into Kenya’s community health strategy could be especially impactful for young mothers. Importantly, the complementary benefits of mHealth and outreach demonstrate that siloed interventions may be less effective than integrated strategies, especially for young mothers facing multi-level barriers to care.

Skilled delivery outcomes further illustrate this point. While only about 4% of participants delivered outside a facility, unskilled deliveries were significantly less common in the mHealth and outreach arms than in the control arm. This finding aligns with evidence from global and Sub-Saharan African studies showing that mHealth interventions, particularly those delivering SMS reminders and education, increase facility-based delivery (21,30,44,49). However, systematic reviews caution that the evidence remains mixed, especially for expectant adolescents and young women (50). Outreach likely amplified these gains by addressing structural barriers such as distance, stigma, sociocultural norms that mHealth alone cannot overcome. Evidence from Tanzania (32) and Kenya (51) suggests that embedding community outreaches within the health system and strengthening community health units could sustainably reduce unskilled deliveries. These findings point to the importance of hybrid interventions, where technology supports demand generation while outreach ensures accessibility and cultural sensitivity.

Low birthweight (<2500 grams) findings revealed a more complex and unexpected picture. While mHealth interventions included nutrition-focused messages, infants in the control group had significantly lower odds of low birthweights compared to intervention arms. This contrasts with other studies that have linked text message-based maternal health interventions to a reduced risk of low birthweight (52–54). Several explanations are possible: biological factors such as maternal height or gestational weight gain, unmeasured in this study, may have confounded the results (55,56). Alternatively, exposure to interventions may not have translated into behavior change if young mothers faced food insecurity, limited autonomy, or lacked resources to act on nutritional advice. This finding reflects inconsistencies noted in systematic reviews, where mHealth showed positive but not always statistically significant effects on low birthweights (57). The association between nulliparity and low birthweight further aligns with evidence linking physiological immaturity to poor birth outcomes (58,59). Also, the finding that participants who received ANC information after pregnancy onset had higher odds of low birthweights also raises questions about the quality, timing, and applicability of information provided. Together, these findings highlight the need for mixed-methods research to unpack how informational interventions intersect with socioeconomic and biological determinants of neonatal outcomes.

From a policy perspective, the results suggest that strengthening maternal outcomes for young mothers requires layered strategies. Technology-driven interventions can boost demand, but without supportive community systems and attention to structural inequities, their impact will be partial. Future programming should consider hybrid models that combine digital health innovations with outreach, health system strengthening, and culturally sensitive engagement.

## Study limitations

This study was not without limitations. As generally expected of quasi experimental studies where participants have not been randomly assigned to conditions as in this study, we observed differences in most of the background characteristics across the three groups. However, significance test of baseline characteristic does not provide an appropriate criterion to assess the effect of imbalance on outcome or the decision to adjust for baseline variables (60,61). This is typical of non-equivalent groups’ studies under the quasi-experimental design where the resulting groups are likely to be dissimilar in some ways. To address potential biases arising from these imbalances, we employed multivariate regression models to adjust for key background characteristics that were likely to influence the outcomes of interest. Another limitation is that this study was conducted in four out of the eleven health centers in Kwale County providing maternal and child health care, findings in this study are therefore not representative of Kwale and hence limited in the extent to which they can be generalized. Despite these limitations, the study provides valuable insights into the effectiveness of mHealth and clinical outreach interventions in improving maternal health outcomes among young women in Kenya.

## Conclusion

This study demonstrates that mHealth text messaging interventions and community-based clinical outreaches can enhance maternal health-seeking behaviors among expectant adolescents and young women, including increased antenatal contacts and skilled deliveries. Combining mHealth with community-based outreaches offers a synergistic approach, leveraging the strengths of both models to improve maternal health outcomes among young women. Findings highlight the need for policy makers and researchers to explore integrated approaches in rural and semi-urban settings across Kenya and sub-Saharan Africa. Certainly, the study’s unexpected findings on infant birthweights underscore the necessity for further research to reconcile inconsistencies with existing evidence. Additionally, future research should explore the combined effects of mHealth and community-based interventions on maternal and neonatal outcomes, assess long-term maternal and child outcomes, and compare integrated versus siloed approaches. Studies should also evaluate cost-effectiveness over extended periods, investigate unmeasured factors influencing birthweight through qualitative and economic analyses, and conduct implementation research to determine scalability in low-resource settings.

## Supporting information

S1 Table. Background characteristics of study participants

S2 Table. A comparison distribution of study participants by their intervention

S3 Table. Effect of mHealth text message and outreach interventions on ANC contacts amongst study participants

S4 Table. Effect of text messaging and outreach interventions on skilled deliveries

S5 Table. Effect of text messaging and outreach interventions on low (<2500) birthweight (<2500) amongst the study participants

## Data Availability

All data produced in the present study are available upon reasonable request to the authors

## Acknowledgments

We extend our sincere gratitude to the Kwale County Department of Health for granting permission to conduct this study within its public health facilities. Special appreciation goes to the in-charges at the study sites for their invaluable support and cooperation throughout the research process. We also acknowledge the dedication and perseverance of our research assistants, whose hard work and patience during data collection, despite the challenges posed by the COVID-19 pandemic were instrumental in the successful completion of this study. Most importantly, we are deeply grateful to all the study participants for their time and willingness to share their experiences, which made this research possible.

## Supporting Information

S1 Table. Background characteristics of study participants

S2 Table. A comparison distribution of study participants by their intervention

S4 Table. Effect of text messaging and outreach interventions on skilled deliveries

## References

1. WHO. WHO Factsheets. 2022 [cited 2022 Feb 11]. Adolescent Pregnancy. Available from: https://www.who.int/en/news-room/fact-sheets/detail/adolescent-pregnancy

2. UNICEF. Monitoring the situation of children and women. 2021 [cited 2021 Feb 7]. Early Child Bearing. Available from: https://data.unicef.org/topic/child-health/adolescent-health/

3. KNBS. Kenya Demographic and Health Survey 2022. Demographic and Health Survey 2022.

4. Mekonnen T, Dune T, Perz J. Maternal health service utilisation of adolescent women in sub-Saharan Africa: A systematic scoping review. BMC Pregnancy Childbirth. 2019;19(1).

5. Tolossa T, Gold L, Lau EH, Dheresa M, Abimanyi-Ochom J. Association between quality of antenatal care service utilisation and adverse birth outcomes among adolescent women in 22 Sub-Saharan African countries. A mixed-effects multilevel analysis. Sex Reprod Healthc. 2024;42.

6. Nderitu CM, Wanyoike-Gichuhi J, Ondieki DK, Odawa X. Pregnancy outcome among adolescents and non-adolescents delivering at Kiambu County Hospital, Kenya. East Afr Med J. 2015;92(8):381–8.

7. Lateef MA, Kuupiel D, Mchunu GG, Pillay JD. Utilization of Antenatal Care and Skilled Birth Delivery Services in Sub-Saharan Africa: A Systematic Scoping Review. Vol. 21, International Journal of Environmental Research and Public Health. 2024.

8. Kuhnt J, Vollmer S. Antenatal care services and its implications for vital and health outcomes of children: Evidence from 193 surveys in 69 low-income and middle-income countries. BMJ Open. 2017;7(11):1–7.

9. Arunda M, Emmelin A, Asamoah BO. Effectiveness of antenatal care services in reducing neonatal mortality in Kenya: Analysis of national survey data. Glob Health Action 2017;10(1). Available from: 10.1080/16549716.2017.1328796

10. Mwaniki MK, Vaid S, Chome IM, Amolo D, Tawfik Y, Coaches KI, et al. Improving service uptake and quality of care of integrated maternal health services: The Kenya kwale district improvement collaborative. BMC Health Serv Res. 2014;14(1):1–9. Available from: https://bmchealthservres.biomedcentral.com/articles/10.1186/1472-6963-14-416

11. Gitobu CM, Gichangi PB, Mwanda WO. The effect of Kenya’s free maternal health care policy on the utilization of health facility delivery services and maternal and neonatal mortality in public health facilities. BMC Pregnancy Childbirth. 2018;18(1):1–11. Available from: https://www.ncbi.nlm.nih.gov/pmc/articles/PMC5870237/pdf/12884_2018_Article_1708.pdf

12. Ndambuki SM, Oyindamola YB, Aimakhu CO. Factors Influencing Utilization of Antenatal Care Services Among Teenage Mothers in Malindi Sub-County Kenya-A Cross Sectional Study. Sci J Public Heal. 2017;5(2):61. Available from: http://www.sciencepublishinggroup.com/journal/paperinfo?journalid=251&doi=10.11648/j.sjph.20170502.12

13. Turyasiima M, Tugume R, Openy A, Ahairwomugisha E, Opio R, Ntuguka M, et al. Determinants of first antenatal care visit by pregnant women at community based education, research and service sites in Northern Uganda. East Africa Med J. 2015;91(9):317–22. Available from: https://www.ncbi.nlm.nih.gov/pmc/articles/PMC4667733/pdf/nihms715819.pdf

14. WHO. WHO recommendations on antenatal care for a positive pregnancy experience Vol. 91, WHO press. Geneva; 2016. Available from: www.who.int/reproductivehealth/publications/maternal_perinatal_health/anc-positive-pregnancy-experience/en/

15. Mash B, Ray S, Essuman A, Burgueño E. Community-orientated primary care: a scoping review of different models, and their effectiveness and feasibility in sub-Saharan Africa. BMJ Glob Heal. 2019;4(Suppl 8):e001489.

16. Annobil I, Dakyaga F, Sillim ML. “From experts to locals hands” healthcare service planning in sub-Saharan Africa: an insight from the integrated community case management of Ghana. BMC Health Serv Res. 2021;21(1):1–15.

17. Ministry of Health Kenya. Kenya Community Health Strategy 2020 - 2025. Nairobi; 2020. Available from: https://www.health.go.ke/wp-content/uploads/2021/01/Kenya-Community-Health-Strategy-Final-Signed-off_2020-25.pdf

18. Endehabtu B, Weldeab A, Were M, Lester R, Worku A, Tilahun B. Mobile phone access and willingness among mothers to receive a text-based mhealth intervention to improve prenatal care in northwest ethiopia: Cross-sectional study. JMIR Pediatr Parent. 2018;1(2):1–13.

19. Fedele DA, Cushing CC, Fritz A, Amaro CM, Ortega A. Mobile health interventions for improving health outcomes in youth a meta-analysis. JAMA Pediatr. 2017;171(5):461–9. Available from: https://www.ncbi.nlm.nih.gov/pmc/articles/PMC6037338/

20. Coleman J, Black V, Thorson AE, Eriksen J. Evaluating the effect of maternal mHealth text messages on uptake of maternal and child health care services in South Africa: a multicentre cohort intervention study. Reprod Health. 2020;17(1):1–9.

21. Shiferaw S, Spigt M, Tekie M, Abdullah M, Fantahun M, Dinant GJ. The effects of a locally developed mHealth intervention on delivery and postnatal care utilization; A prospective controlled evaluation among health centres in Ethiopia. PLoS One. 2016;11(7):1–14.

22. Colaci D, Chaudhri S, Vasan A. mHealth Interventions in Low-Income Countries to Address Maternal Health_: A Systematic Review. Ann Glob Heal. 2016;82(5):922–35.

23. Sewpaul R, Resnicow K, Crutzen R, Dukhi N, Ellahebokus A, Reddy P. A Tailored mHealth Intervention for Improving Antenatal Care Seeking and Health Behavioral Determinants During Pregnancy Among Adolescent Girls and Young Women in South Africa: Development and Protocol for a Pilot Randomized Controlled Trial. JMIR Res Protoc. 2023;12(December 2019):1–14.

24. Benski AC, Schmidt NC, Viviano M, Stancanelli G, Soaroby A, Reich MR. Improving the quality of antenatal care using mobile health in madagascar: Five-year cross-sectional study. JMIR mHealth uHealth. 2020;8(7).

25. 25. Ippoliti N. Mobile Phone Programs For Adolescent And Youth Sexual And Reproductive Health In Low- And Middle- Income Countries. 2017. Available from: https://www.k4health.org/sites/default/files/youth-mhealth-srh-brief-2.pdf

26. Lund S, Nielsen BB, Hemed M, Boas IM, Said A, Said K, et al. Mobile phones improve antenatal care attendance in Zanzibar: A cluster randomized controlled trial. BMC Pregnancy Childbirth 2014;14(1):1–10. Available from: https://bmcpregnancychildbirth.biomedcentral.com/articles/10.1186/1471-2393-14-29

27. Ridgeway K, Dulli LS, Murray KR, Silverstein H, Santo LD, Olsen P, et al. Interventions to improve antiretroviral therapy adherence among adolescents in low- and middle-income countries: A systematic review of the literature. Vol. 13, PLoS ONE. 2018. 1–33 p.

28. Watterson JL, Walsh J, Madeka I. Using mHealth to Improve Usage of Antenatal Care, Postnatal Care, and Immunization: A Systematic Review of the Literature. Biomed Res Int 2015;2015. Available from: https://www.ncbi.nlm.nih.gov/pubmed/26380263

29. Lee S, Ulugbek B, Mukherjee M, Grant L, Pagliari C. Effectiveness of mHealth interventions for maternal, newborn and child health in low – and middle – income countries_: Systematic review and meta – analysis. Glob Heal. 2016;6(1). Available from: https://www.ncbi.nlm.nih.gov/pmc/articles/PMC4643860/

30. Maliwichi P, Chigona W, Sowon K. Appropriation of mHealth interventions for maternal health care in Sub-Saharan Africa: Hermeneutic review. JMIR mHealth uHealth. 2021;9(10).

31. Bang K sook, Chae S mi, Lee I, Yu J, Kim J. Effects of a Community Outreach Program for Maternal Health and Family Planning in Tigray, Ethiopia. Asia Nurs Res. 2018;12.

32. Geldsetzer P, Mboggo E, Larson E, Lema IA, Magesa L, Machumi L, et al. Community health workers to improve uptake of maternal healthcare services: A cluster randomized pragmatic trial in dar es salaam, tanzania. PLoS Med. 2019;16(3):1–27.

33. Ronen K, McGrath CJ, Langat AC, Kinuthia J, Omolo D, Singa B, et al. Gaps in Adolescent Engagement in Antenatal Care and Prevention of Mother-to-Child HIV Transmission Services in Kenya. J Acquir Immune Defic Syndr. 2017;74(1):30–7. Available from: https://www.ncbi.nlm.nih.gov/pmc/articles/PMC5895459/pdf/nihms954809.pdf

34. 34. Babycentre. Mission Motherhood Messages. 2017 [cited 2020 Jan 8]. Available from: https://www.babycenter.com/mission-motherhood/messages/

35. 35. UNICEF. Maternal and Newborn Health Disparities. Nairobi; 2018. Available from: https://data.unicef.org/resources/maternal-newborn-health-disparities-country-profiles/

36. 36. Ministry of Health Kenya. Kenya Reproductive, Maternal, Newborn, Child and Adolescent Health (RMNCAH) Investment Framework Nairobi; 2016. Available from: https://www.globalfinancingfacility.org/sites/gff_new/files/Kenya-Investment-Case.pdf

37. Ikamari L. Uptake of maternal services and associated factors in the western region of Kenya. Pan Afr Med J. 2020;37(192):1–15.

38. Anaba EA, Alor SK, Badzi CD. Utilization of antenatal care among adolescent and young mothers in Ghana; analysis of the 2017/2018 multiple indicator cluster survey. BMC Pregnancy Childbirth. 2022;22(1):1–8. Available from: 10.1186/s12884-022-04872-z

39. Hackett K, Lenters L, Vandermorris A, Lafleur C, Newton S, Ndeki S, et al. How can engagement of adolescents in antenatal care be enhanced? Learning from the perspectives of young mothers in Ghana and Tanzania. BMC Pregnancy Childbirth. 2019;19(1):1–12.

40. Okedo-Alex IN, Akamike IC, Ezeanosike OB, Uneke CJ. Determinants of antenatal care utilisation in sub-Saharan Africa: A systematic review. BMJ Open. 2019;9(10).

41. Ng’ambi W, Collins J, Colbourn T, Mangal T, Phillips A, Kachale F, et al. Socio- demographic factors associated with early antenatal care visits among pregnant women in Malawi: 2004-2016. medRxiv. 2021;2021.10.08.21264750. Available from: http://medrxiv.org/content/early/2021/10/11/2021.10.08.21264750.abstract

42. Amo-Adjei J, Anamaale Tuoyire D. Effects of planned, mistimed and unwanted pregnancies on the use of prenatal health services in sub-Saharan Africa: a multicountry analysis of Demographic and Health Survey data. Trop Med Int Heal. 2016;21(12):1552–61.

43. Felisian S, Mushy SE, Tarimo EAM, Kibusi SM. Sociocultural practices and beliefs during pregnancy, childbirth, and postpartum among indigenous pastoralist women of reproductive age in Manyara, Tanzania: a descriptive qualitative study. BMC Womens Health. 2023;23(1):1–8.

44. Kabongo EM, Mukumbang FC, Delobelle P, Nicol E. Explaining the impact of mHealth on maternal and child health care in low- and middle-income countries_: a realist synthesis. BMC Pregnancy Childbirth. 2021;21(196).

45. Dol J, Richardson B, Tomblin Murphy G, Aston M, McMillan D, Campbell-Yeo M. Impact of mobile health (mHealth) interventions during the perinatal period for mothers in low- and middle-income countries: A systematic review. JBI Database Syst Rev Implement Reports. 2019;17(8):1634–67.

46. Arnaert A, Ponzoni N, Debe Z, Meda MM, Nana NG, Arnaert S. Experiences of women receiving mhealth-supported antenatal care in the village from community health workers in rural Burkina Faso, Africa. Digit Heal. 2019;5:1–8. Available from: 10.1177/2055207619892756

47. Shin HY, Kim KY, Kang P. Concept analysis of community health outreach. BMC Health Serv Res. 2020;20(417).

48. Anderson T, Jacqueline R, Brian G, Chelsey B. House Parties_: An Innovative Model for Outreach and Community- Based Health Education. Matern Child Health J. 2017;21(1):75–80.

49. Chen H, Chai Y, Dong L, Niu W, Zhang P. Effectiveness and appropriateness of mhealth interventions for maternal and child health: Systematic review. JMIR mHealth uHealth. 2018;6(1):1–12.

50. Marcolino MS, Oliveira JAQ, D’Agostino M, Ribeiro AL, Alkmim MBM, Novillo-Ortiz D. The impact of mHealth interventions: Systematic review of systematic reviews. J Med Res. 2018;20(1).

51. Kumar MB, Madan JJ, Auguste P, Taegtmeyer M, Otiso L, Ochieng CB, et al. Cost-effectiveness of community health systems strengthening: Quality improvement interventions at community level to realise maternal and child health gains in Kenya. BMJ Glob Heal. 2021;6(3):1–10.

52. Coleman J, Bohlin KC, Thorson A, Black V, Mechael P, Mangxaba J, et al. Effectiveness of an SMS-based maternal mHealth intervention to improve clinical outcomes of HIV- positive pregnant women. AIDS Care. 2017;0(0):1–8.

53. Saronga N, Burrows T, Collins C, Ashman A, Rollo M. mHealth interventions of improving nutrients intake of pregnant women in low and lower-middle income countries: systematic review. Matern Child Nutr. 2019;15.

54. Fedha T. Impact of Mobile Telephone on Maternal Health Service Care: A Case of Njoro Division. Open J Prev Med. 2014;4(4):365–76. Available from: http://www.scirp.org/journal/ojpm%0Ahttp://dx.doi.org/10.4236/ojpm.2014.45044%0Ahttp://creativecommons.org/licenses/by/4.0/

55. Girma S, Fikadu T, Agdew E, Haftu D, Gedamu G, Dewana Z, et al. Factors associated with low birthweight among newborns delivered at public health facilities of Nekemte town, West Ethiopia: A case control study. BMC Pregnancy Childbirth. 2019;19(1):1–6.

56. Desta SA, Damte A, Hailu T. Maternal factors associated with low birth weight in public hospitals of Mekelle city, Ethiopia: A case-control study. Ital J Pediatr. 2020;46(1):1–9.

57. Eberle C, Loehnert M, Stichling S. Effectiveness of specific mobile health applications (mHealth-apps) in gestational diabtetes mellitus_: a systematic review. BMC Pregnancy Childbirth 2021;(2021):1–7. Available from: 10.1186/s12884-021-04274-7

58. Garces A, Perez W, Harrison MS, Hwang KS, Nolen TL, Goldenberg RL, et al. Association of parity with birthweight and neonatal death in five sites: The Global Network’s Maternal Newborn Health Registry study. Reprod Health. 2020;17(3):1–8. Available from: 10.1186/s12978-020-01025-3

59. Nyamasege CK, E.W KM, Wanjohi M, Kaindi DWM, Ma E, Fukushige M, et al. Determinants of low birth weight in the context of maternal nutrition education in urban informal settlements, Kenya. J Dev Orig Heal Dis. 2018;

60. Roberts C, Torgerson DJ. Understanding controlled trials. Baseline imbalance in randomised controlled trials. Br Med J. 1999;319(7203):185. Available from: 10.1136/bmj.319.7203.185

61. Senn SJ. Covariate imbalance and random allocation in clinical trials. Stat Med. 1989;8(4):467–75. Available from: doi: 10.1002/sim.4780080410. PMID: 2727470

