## Supplementary material for "Improving maternal outcomes of young mothers through mobile health (mHealth) and community-based interventions: Evidence from a quasi-experimental trial in Kwale County, Coastal Kenya": S1 Table. Background characteristics of study participants

Supplemental table 1: Background characteristics of study participants (N=817)

|  | **Mkongani**  **(n=213)** | **Kikoneni**  **(n=205)** | **Mnyenzeni**  **(n=193)** | **Mazeras**  **(n=206)** | **Overall**  **(n=817)** |
| --- | --- | --- | --- | --- | --- |
| *Mean age (standard deviation)* | 20.4 (2.4) | 20.8 (2.6) | 21.7 (2.4) | 21.4 (2.0) | 21.1 (2.4) |
| *Age group (years):*  15-19  20-24 | 73 (34.3)  140 (65.7) | 66 (32.2)  139 (67.8) | 37 (19.2)  156 (80.8) | 39 (18.9)  167 (81.1) | 215 (26.3)  602 (73.7) |
| *Highest education level:*  Incomplete primary  Complete primary  Secondary and above  Missing | 107 (50.2)  76 (35.7)  30 (14.1)  0 (0.0) | 104 (50.7)  70 (34.2)  31 (15.1)  0 (0.0) | 106 (54.9)  59 (30.6)  26 (13.5)  2 (1.0) | 52 (25.2)  98 (47.6)  56 (27.2)  0 (0.0) | 369 (45.2)  303 (37.1)  143 (17.5)  2 (0.2) |
| *Religion:*  Christian  Muslim  Others | 43 (20.2)  170 (79.8)  0 (0.0) | 31 (15.1)  174 (84.9)  0 (0.0) | 53 (27.5)  135 (70.0)  5 (2.6) | 103 (50.0)  102 (49.5)  1 (0.5) | 230 (28.2)  581 (71.1)  6 (0.7) |
| *Mean age at sex debut (sd)* | 17.4 (1.8) | 17.2 (2.2) | 15.9 (2.0) | 19.0 (1.6) | 17.4 (2.2) |
| *Age at sex debut:*  10-14 years  15-17 years  18-24 years  Missing | 8 (3.8)  95 (44.6)  110 (51.6)  0 (0.0) | 17 (8.3)  91 (44.4)  82 (40.0)  15 (7.3) | 59 (30.6)  84 (43.5)  50 (25.9)  0 (0.0) | 3 (1.5)  18 (8.7)  177 (85.9)  8 (3.9) | 87 (10.7)  288 (35.2)  419 (51.3)  23 (2.8) |
| *Marital status:*  Single /Dating/Cohabiting  Engaged/Married | 29 (13.6)  184 (86.4) | 76 (37.1)  129 (62.9) | 5 (2.6)  188 (97.4) | 23 (11.2)  183 (88.8) | 133 (16.3)  684 (83.7) |
| *Primary caregiver:*  None  Male partner  Parents  Siblings/Relatives/Others | 2 (0.9)  182 (85.5)  26 (12.2)  3 (1.4) | 1 (0.5)  105 (51.2)  91 (44.4)  8 (3.9) | 2 (1.0)  179 (92.8)  11 (5.7)  1 (0.5) | 1 (0.5)  181 (87.9)  22 (10.7)  2 (1.0) | 6 (0.7)  647 (79.2)  150 (18.4)  14 (1.7) |
| *Mean age of male partner (sd)* | 25.7 (4.1) | 26.9 (6.4) | 28.0 (4.9) | 26.6 (3.4) | 26.8 (4.8) |
| *Age group of male partners:*  17-24 years  25-29 years  30+ years  Missing | 84 (39.4)  101 (47.4)  28 (13.2)  0 (0.0) | 61 (29.8)  74 (36.1)  40 (19.5)  30 (14.6) | 48 (24.9)  76 (39.4)  69 (35.8)  0 (0.0) | 45 (21.8)  108 (52.4)  40 (19.4)  13 (6.3) | 238 (29.1)  359 (43.9)  177 (21.7)  43 (5.3) |
| *Employed or in business:*  No  Yes | 201 (94.4)  12 (5.6) | 183 (89.3)  22 (10.7) | 169 (87.6)  24 (12.4) | 189 (91.8)  17 (8.3) | 742 (90.8)  75 (9.2) |
| *Planned pregnancy:*  Yes  No  Unsure | 112 (52.6)  100 (47.0)  1 (0.5) | 121 (59.0)  78 (38.1)  6 (2.9) | 175 (90.7)  18 (9.3)  0 (0.0) | 180 (87.4)  26 (12.6)  0 (0.0) | 588 (72.0)  222 (27.2)  7 (0.9) |
| *Parity:*  Nulliparous  1  2  3+ | 114 (53.5)  50 (23.5)  42 (19.7)  7 (3.3) | 131 (63.9)  37 (18.1)  21 (10.2)  16 (7.8) | 61 (31.6)  44 (22.8)  36 (18.7)  52 (26.9) | 161 (78.2)  33 (16.0)  5 (2.4)  7 (3.4) | 467 (57.2)  164 (20.1)  104 (12.7)  82 (10.0) |
| *Mean age at first pregnancy (sd)* | 18.5 (2.1) | 19.0 (2.5) | 17.3 (2.3) | 20.5 (1.9) | 18.8 (2.5) |
| *Age group at first pregnancy:*  14 - 17 years  18-19 years  20-24 years | 63 (29.6)  87 (40.9)  63 (29.6) | 60 (29.3)  69 (33.7)  76 (37.1) | 101 (52.3)  53 (27.5)  39 (20.2) | 8 (3.9)  57 (27.7)  141 (68.5) | 232 (28.4)  266 (32.6)  319 (39.1) |
| *Mean Gestational age in weeks at first ANC* | 19.3 (4.4) | 20.5 (3.9) | 18.8 (5.7) | 19.5 (4.6) | 19.6 (4.8) |
| *Gestation at first ANC:*  0-13 weeks  14-19 weeks  20-24 weeks | 21 (9.9)  38 (17.8)  154 (72.3) | 12 (5.9)  45 (22.0)  148 (72.2) | 47 (24.4)  20 (10.4)  126 (65.3) | 19 (9.2)  43 (20.9)  144 (70.0) | 99 (12.1)  146 (17.9)  572 (70.0) |
| *Received any information on the importance of ANC since becoming pregnant:*  No  Yes | 134 (62.9)  79 (37.1) | 63 (30.7)  142 (69.3) | 15 (7.8)  178 (92.2) | 114 (55.3)  92 (44.7) | 326 (40.0)  491 (60.1) |
| *Knowledge on when a woman should start ANC clinic:*  On missing periods  1^st^ trimester  2^nd^ trimester  3^rd^ trimester  Don’t know | 13 (6.1)  28 (13.2)  110 (51.6)  22 (10.3)  40 (18.8) | 21 (10.2)  92 (44.9)  73 (35.6)  3 (1.5)  16 (7.8) | 16 (8.3)  155 (80.3)  21 (10.9)  1 (0.5)  0 (0.0) | 11 (5.3)  63 (30.6)  103 (50.0)  3 (1.5)  26 (12.6) | 61 (7.5)  338 (41.4)  307 (37.6)  29 (3.6)  82 (10.0) |
| *Knowledge on how many ANC visits is recommended:*  *None*  *Between 1-2*  *Between 3-5*  *At least 4*  *As many as possible* | 10 (4.7)  2 (0.9)  21 (9.9)  80 (37.6)  100 (46.9) | 1 (0.5)  0 (0.0)  82 (40.0)  48 (23.4)  74 (36.1) | 0 (0.0)  33 (17.1)  19 (9.8)  73 (37.8)  68 (35.2) | 14 (6.8)  1 (0.5)  77 (37.4)  49 (23.8)  65 (31.6) | 25 (3.1)  36 (4.4)  199 (24.4)  250 (30.6)  307 (37.6) |
| *Time to facility (minutes):*  0-30  31-60  61+ | 53 (24.9)  77 (36.2)  83 (39.0) | 59 (28.8)  79 (38.5)  67 (32.7) | 34 (17.6)  77 (39.9)  82 (42.5) | 95 (46.1)  84 (40.8)  27 (13.1) | 241 (29.5)  317 (38.8)  259 (31.7) |
