## Supplementary material for "Improving maternal outcomes of young mothers through mobile health (mHealth) and community-based interventions: Evidence from a quasi-experimental trial in Kwale County, Coastal Kenya": S2 Table. A comparison distribution of study participants by their intervention

Supplemental table 2: A comparison distribution of study participants by their intervention status (N=817)

| **Characteristics** | **Control**  **(n=272)** | **Intervention**  **(n=398)** | **Outreach**  **(n=147)** | **p-value*** |
| --- | --- | --- | --- | --- |
| *Median age (IQR)* | 21 (20-23) | 21(19-24) | 20 (19-22) | <0.001 |
| *Age group (years):*  15-19  20-24 | 55 (20.2)  217 (79.8) | 103 (25.9)  295 (74.1) | 57 (38.8)  90 (61.2) | <0.001 |
| *Highest education level:*  Incomplete primary  Complete primary  Secondary and above  Missing | 86 (31.6)  118 (43.4)  68 (25.0)  0.0 | 210 (52.8)  129 (32.4)  57 (14.3)  2 (0.5) | 73 (49.7)  56 (38.1)  18 (12.2)  0.0 | <0.001 |
| *Religion:*  Christian  Muslim  Others | 114 (41.9)  157 (57.7)  1 (0.37) | 84 (21.1)  309 (77.6)  5 (1.3) | 32 (21.8)  115 (78.2)  0.0 | <0.001 |
| *Median age at sex debut (IQR)* | 19 (18-20) | 16 (15-18) | 18 (16-18) | 0.0001 |
| *Age group at sexual debut:*  *10-14 years*  *15-17 years*  *18-24 years*  *Missing* | 6 (2.2)  48 (17.7)  210 (77.2)  8 (2.9) | 76 (19.1)  175 (44.0)  132 (33.2)  15 (3.8) | 5 (3.4)  65 (44.2)  77 (52.4)  0.0 | <0.001 |
| *Marital status:*  Single /Dating/Cohabiting  Engaged/Married | 29 (10.7)  243 (89.3) | 81 (20.4)  317 (79.7) | 23 (15.7)  124 (84.4) | 0.004 |
| *Primary caregiver:*  None  Male partner  Parents  Relatives/Neighbors/Others | 2 (0.7)  239 (87.9)  28 (10.3)  3 (1.1) | 3 (0.8)  284 (71.4)  102 (25.6)  9 (2.3) | 1 (0.7)  124 (84.4)  20 (13.6)  2 (1.4) | <0.001 |
| *Media age of male partner (IQR)* | 26 (24-28) | 26 (24-30) | 25 (23-28) | 0.003 |
| *Age group of male partners:*  <25 years  25-29 years  30+ years  Missing | 71 (26.1)  139 (51.1)  49 (18.0)  13 (4.8) | 109 (27.4)  150 (37.7)  109 (27.4)  30 (7.5) | 58 (39.5)  70 (47.6)  19 (12.9)  0.0 | <0.001 |
| *Employed or in business:*  No  Yes | 250 (91.9)  22 (8.1) | 352 (88.4)  6 (11.6) | 140 (95.2)  7 (4.8) | 0.04 |
| *Planned pregnancy:*  Yes  No  Unsure | 220 (80.9)  51 (18.8)  1 (0.4) | 296 (74.4)  96 (24.1)  6 (1.5) | 72 (49.0)  75 (51.0)  0.0 | <0.001 |
| *Parity:*  Nulliparous  1  2  3+ | 195 (71.7)  47 (17.3)  20 (7.4)  10 (3.7) | 192 (48.2)  81 (20.4)  57 (14.3)  68 (17.1) | 80 (54.4)  36 (24.5)  27 (18.4)  4 (2.7) | <0.001 |
| *Median age at first pregnancy (IQR)* | 20 (18-21) | 18 (16-20) | 18 (17-20) | 0.0001 |
| *Age group at first pregnancy:*  14 -17 years  18 – 19 years  20 – 24 years | 32 (11.8)  77 (28.3)  163 (60.0 | 161 (40.5)  122 (30.7)  67 (45.6) | 39 (26.5)  67 (45.6)  41 (27.9) | <0.001 |
| *Gestation at first ANC:*  0-13 weeks  14-19 weeks  20-24 weeks | 26 (9.6)  56 (20.6)  190 (69.9) | 59 (14.8)  65 (16.3)  274 (68.8) | 14 (9.5)  25 (17.0)  108 (73.5) | 0.151 |
| *Received any ANC information since becoming pregnant:*  No  Yes | 147 (54.0)  125 (46.0) | 78 (19.6)  320 (80.4) | 101 (68.7)  46 (31.3) | <0.001 |
| *Knowledge on when a woman should start ANC clinic:*  On missing periods  1^st^ trimester  2^nd^ trimester  3^rd^ trimester  Don’t know | 15 (5.5)  76 (27.9)  138 (50.7)  6 (2.2)  37 (13.6) | 37 (9.3)  247 (62.1)  94 (23.6)  4 (1.0)  16 (4.0) | 9 (6.1)  15 (10.2)  75 (51.0)  19 (12.9)  29 (19.7) | <0.001 |
| *Knowledge on how many ANC visits is recommended:*  *None*  *Between 1-2*  *Between 3-5*  *At least 4*  *As many as possible* | 18 (6.6)  3 (1.1)  83 (30.5)  77 (28.3)  91 (33.5) | 1 (0.3)  33 (8.3)  101 (25.4)  121 (30.4)  142 (35.7) | 6 (4.1)  0.0  15 (10.2)  52 (35.4)  74 (50.3) | <0.001 |
| *Time to facility (mins):*  0-30  31-60  61+ | 115 (42.3)  109 (40.1)  48 (17.7) | 93 (23.4)  156 (39.2)  149 (37.4) | 33 (22.5)  52 (35.4)  62 (42.2) | <0.001 |
| *Continuous variables assessed using Kruskal-Wallis test; categorical variables assessed using the chi-square test | | | | |
