## Supplementary material for "Improving maternal outcomes of young mothers through mobile health (mHealth) and community-based interventions: Evidence from a quasi-experimental trial in Kwale County, Coastal Kenya": S3 Table. Effect of mHealth text message and outreach interventions on ANC contacts amongst study participants

Supplemental table 3: Effect of mHealth text message and outreach interventions on ANC contacts amongst study participants (N=817)

| **Characteristics** | **Low ANC visits (%)** | **Crude Risk Ratio**  **(95% CI)** | **p-value** | **Adjusted Risk Ratio**  **(95% CI)** | **p-value** |
| --- | --- | --- | --- | --- | --- |
| *Interventions:*  Control  MHealth intervention  Outreach participants | 150 (55.1)  119 (29.0)  4 (2.7) | Ref  0.5 (0.4-0.6)  0.04 (0.02-0.1) | <0.001  <0.001 | Ref  0.5 (0.4 – 0.7)  0.1 (0.0 – 0.7) | <0.001  0.024 |
| *Age group (years):*  15-19  20-24 | 59 (27.4)  214 (35.6) | Ref  1.3 (1.0-1.7) | 0.04 | Ref  0.7 (0.5 – 1.0) | 0.063 |
| *Highest education level:*  Incomplete primary  Complete primary  Secondary and above | 108 (29.3)  107 (35.3)  56 (39.2) | Ref  1.2 (1.0 – 1.5)  1.3 (1.0 – 1.7) | 0.094  0.027 | Ref  1.1 (0.9 – 1.3)  1.2 (1.0 – 1.5) | 0.382  0.080 |
| *Religion:*  Christian  Muslim  Others | 104 (45.2)  166 (28.6)  3 (50.0) | Ref  0.6 (0.5 – 0.8)  1.1 (0.5 – 2.5) | <0.001  0.808 | 0.9 (0.8 -1.1)  0.8 (0.4 – 1.9) | 0.410  0.660 |
| *Age group at sexual debut:*  *10-14 years*  *15-17 years*  *18-24 years* | 32 (36.8)  66 (22.9)  160 (38.2) | Ref  0.6 (0.4 – 0.9)  1.0 (0.8 – 1.4) | 0.008  0.807 | Ref  0.7 (0.5 – 1.1)  0.7 (0.5 – 1.1) | 0.094  0.094 |
| *Marital status:*  Single /Dating/Cohabiting  Engaged/Married | 30 (22.6)  243 (35.5) | Ref  1.6 (1.1 – 2.2) | 0.007 | 1.2 (0.9 -1.6) | 0.305 |
| *Primary caregiver:*  None  Male partner  Parents  Relatives/Neighbors/Others | 2 (33.3)  240 (37.1)  28 (18.7)  3 (21.4) | Ref  1.1 (0.4 – 3.5)  0.6 (0.2 – 1.8)  0.6 (0.1 – 2.9) | 0.854  0.335  0.567 |  |  |
| *Age group of male partners:*  <25 years  25-29 years  30+ years | 65 (27.3)  123 (34.3)  68 (38.4) | Ref  1.3 (1.0 – 1.6)  1.4 (1.1 – 1.9) | 0.078  0.016 | 1.1 (0.9 – 1.4)  1.3 (1.0 – 1.6) | 0.362  0.080 |
| *Employed or in business:*  No  Yes | 247 (33.3)  26 (34.7) | Ref  1.0 (0.8 – 1.4) | 0.808 |  |  |
| *Planned pregnancy:*  No  Yes | 32 (14.4)  241 (41.0) | Ref  2.8 (2.0 – 4.0) | <0.001 | Ref  2.3 (1.5 – 3.5) | <0.001 |
| *Parity:*  Nulliparous  1  2  3+ | 169 (36.2)  52 (31.7)  25 (24.0)  27 (32.9) | Ref  0.9 (0.7 – 1.1)  0.7 (0.5 – 1.0)  0.9 (0.7 – 1.3) | 0.309  0.027  0.577 | Ref  1.1 (0.8 – 1.4)  0.7 (0.5 – 1.1)  0.8 (0.6 – 1.3) | 0.572  0.128  0.399 |
| *Age group at first pregnancy:*  14 - 17 years  18 – 19 years  20 – 24 years | 63 (27.2)  77 (29.0)  133 (41.7) | Ref  1.1(0.8 – 1.4)  1.5 (1.2 – 2.0) | 0.658  0.001 | Ref  1.1 (0.8 – 1.5)  1.2 (0.8 – 1.8) | 0.621  0.383 |
| *Gestation at first ANC:*  0-13 weeks  14-19 weeks  20-24 weeks | 24 (24.2)  41 (28.1)  208 (36.4) | Ref  1.2 (0.8 – 1.8)  1.5 (1.0 – 2.2) | 0.507  0.029 | Ref  1.2 (0.9 – 1.7)  1.6 (1.2 – 2.1) | 0.257  0.003 |
| *Received any ANC information since becoming pregnant:*  No  Yes | 92 (28.2)  181 (36.9) | Ref  1.3 (1.1 – 1.6) | 0.012 | Ref  1.0 (0.8 – 1.2) | 0.772 |
| *Knowledge on when to start ANC clinic:*  On missing periods  1^st^ trimester  2^nd^ trimester  3^rd^ trimester  Don’t know | 16 (26.2)  139 (41.1)  102 (33.2)  2 (6.9)  14 (17.1) | Ref  1.6 (1.0 – 2.4)  1.3 (0.9 – 2.0)  0.3 (0.1 – 1.2)  0.7 (0.3 – 1.2) | 0.045  0.303  0.062  0.186 | Ref  1.4 (1.0 – 2.1)  1.1 (0.7 – 1.6)  0.2 (0.1 – 3.2)  1.0 (0.4 – 1.4) | 0.053  0.771  0.242  0.362 |
| *Knowledge on how many ANC visits is recommended:*  *None*  *Between 1-2*  *Between 3-5*  *At least 4*  *As many as possible* | 7 (28.0)  22 (61.1)  79 (39.7)  92 (36.8)  73 (23.8) | Ref  2.2 (1.2 – 4.3)  1.4 (0.7 – 2.7)  1.3 (0.7 – 2.5)  0.8 (0.4 – 1.6) | 0.025  0.294  0.409  0.627 | Ref  1.8 (1.0 – 3.5)  1.0 (0.6 – 1.9)  1.2 (0.7 – 2.1)  0.9 (0.5 – 1.6) | 0.061  0.880  0.609  0.667 |
| *Time to facility (minutes):*  0-30  31-60  61+ | 80 (33.2)  114 (36.0)  79 (30.5) | Ref  1.1 (0.9 – 1.4)  0.9 (0.7 – 1.2) | 0.498  0.518 |  |  |

*ANC = Antenatal Care; Adjusted analyses include age, education, religion, age at sex debut, marital status, planned pregnancy, parity, and time to facility
