## Supplementary material for "Improving maternal outcomes of young mothers through mobile health (mHealth) and community-based interventions: Evidence from a quasi-experimental trial in Kwale County, Coastal Kenya": S4 Table. Effect of text messaging and outreach interventions on skilled deliveries

Supplemental table 4: Effect of text messaging and outreach interventions on skilled deliveries

| **Characteristics** | **TBA (%)** | **Crude Odd Ratio**  **(95% CI)** | **p-value** | **Adjusted Odd Ratio**  **(95% CI)** | **p-value** |
| --- | --- | --- | --- | --- | --- |
| *Interventions:*  Control  MHealth intervention  Outreach participants | 24 (8.8)  5 (1.3)  2 (1.4) | Ref  0.1 (0.04 – 0.3)  0.1 (0.03 – 0.6) | <0.001  0.08 | Ref  0.2 (0.1 – 0.4)  0.1 (0.0 – 0.6) | <0.001  0.007 |
| *Age group (years):*  15-19  20-24 | 6 (2.8)  25 (4.2) | Ref  1.5 (0.6 – 3.7) | 0.376 |  |  |
| *Highest education level:*  Incomplete primary  Complete primary  Secondary and above | 6 (1.6)  19 (6.3)  6 (4.2) | Ref  4.1 (1.6 – 10.5)  2.7 (0.8 – 8.4) | 0.003  0.094 | Ref  3.2 (1.2 – 8.2)  1.5 (0.5 – 5.0) | 0.019  0.472 |
| *Religion:*  Christian  Muslim  Others | 12 (5.2)  18 (3.1)  1 (16.7) | Ref  0.6 (0.3 – 1.2)  3.5 (0.4 – 32.0) | 0.135  0.275 |  |  |
| *Age at sexual debut:*  *10-14 years*  *15-17 years*  *18-24 years* | 2 (2.3)  4 (1.4)  25 (5.7) | Ref  0.6 (0.1 – 3.2)  2.6 (0.6 – 11.0) | 0.535  0.208 |  |  |
| *Marital status:*  Single /Dating/Cohabiting  Engaged/Married | 2 (1.5)  29 (4.2) | Ref  2.8 (0.7 – 12.1) | 0.156 |  |  |
| *Primary caregiver:*  None  Male partner  Parents  Relatives/Neighbors | 0 (0.0)  29 (4.5)  2 (1.3)  0 (0.0) | Ref  0.1 (0.0 – 0.8) | 0.096 |  |  |
| *Age group of male partners:* 17-24 years  25-29 years  30+ years  Missing | 6 (2.5)  17 (4.7)  5 (2.8)  3 (7.0) | Ref  1.9 (0.7 – 4.8)  1.1 (0.3 – 3.6) | 0.200  0.887 |  |  |
| *Employed or in business:*  No  Yes | 30 (4.0)  1 (1.3) | Ref  0.3 (0.04 – 2.3) | 0.258 |  |  |
| *Planned pregnancy:*  No  Yes | 6 (2.7)  24 (4.1) | Ref  1.6 (0.6 – 3.8) | 0.343 |  |  |
| *Parity:*  Nulliparous  1  2  3+ | 16 (3.4)  7 (4.3)  5 (4.8)  3 (3.7) | Ref  1.2 (0.5 – 3.1)  1.4 (0.5 – 3.9)  1.0 (0.3 – 3.7) | 0.644  0.526  0.950 |  |  |
| *Age group at first pregnancy:*  14 – 17 years  18 – 19 years  20 – 24 years | 6 (2.6)  11 (4.1)  14 (4.4) | Ref  1.6 (0.6 – 4.5)  1.8 (0.7 – 4.7) | 0.341  0.252 |  |  |
| *Gestation at first ANC:*  0-13 weeks  14-19 weeks  20-24 weeks | 5 (5.1)  4 (2.7)  22 (3.9) | Ref  0.5 (0.1 – 2.0)  0.8 (0.3 – 2.0) | 0.356  0.574 |  |  |
| *Received any ANC information since becoming pregnant:*  No  Yes | 17 (5.2)  14 (2.9) | Ref  0.5 (0.3 – 1.1) | 0.093 |  |  |
| *Knowledge on when to start ANC clinic:*  On missing periods  1^st^ trimester  2^nd^ trimester  3^rd^ trimester  Don’t know | 4 (7.1)  9 (2.7)  14 (4.7)  1 (3.6)  3 (3.9) | Ref  0.4 (0.1 – 1.2)  0.6 (0.2 – 2.0)  0.5 (0.1 – 4.5)  0.5 (0.1 – 2.4) | 0.103  0.441  0.523  0.405 |  |  |
| *Knowledge on how many ANC visits is recommended:*  *None*  *Between 1-2*  *Between 3-5*  *At least 4*  *As many as possible* | 3 (12.0)  0 (0.0)  5 (2.5)  9 (3.6)  14 (4.6) | Ref  (empty)  0.2 (0.0 – 0.8)  0.3 (0.6 – 1.0)  0.3 (0.1 – 1.2) | 0.024  0.052  0.102 | 0.2 (0.0 – 1.1)  0.5 (0.1 – 2.1)  0.6 (0.1 – 2.4) | 0.072  0.333  0.460 |
| *Time to facility (minutes):*  0-30  31-60  61+ | 14 (5.8)  9 (2.8)  8 (3.1) | Ref  0.5 (0.2 – 1.1)  0.5 (0.2 – 1.2) | 0.078  0.130 |  |  |

*ANC = Antenatal Care; Adjusted analyses include age, education, religion, age at sex debut, marital status, planned pregnancy, parity, and time to facility
