## Supplementary material for "Improving maternal outcomes of young mothers through mobile health (mHealth) and community-based interventions: Evidence from a quasi-experimental trial in Kwale County, Coastal Kenya": S5 Table. Effect of text messaging and outreach interventions on low (<2500) birthweight (<2500) amongst the study participants

Supplemental table 5: Effect of text messaging and outreach interventions on low (<2500) birthweight (<2500) amongst the study participants

| **Characteristics** | **Low birth weight (%)** | **Crude Odd Ratio**  **(95% CI)** | **p-value** | **Adjusted Odd Ratio**  **(95% CI)** | **p-value** |
| --- | --- | --- | --- | --- | --- |
| *Interventions:*  *Pure control*  *MHealth intervention*  *Outreach participants* | 3 (1.2)  39 (10.1)  9 (6.2) | Ref  9.6 (2.9 – 39.4)  5.6 (1.5 – 21.2) | <0.001  0.010 | Ref  5.9 (1.7 – 20.8)  6.6 (1.7 – 25.7) | 0.006  0.007 |
| *Age group (years):*  15 – 19  20 - 25 | 11 (5.3)  40 (6.9) | Ref  1.3 (0.7 – 2.6) | 0.435 |  |  |
| *Highest education level:*  Incomplete primary  Complete primary  Secondary and above | 21 (5.8)  19 (6.6)  10 (7.2) | Ref  1.1 (0.6 – 2.2)  1.3 (0.6 – 2.8) | 0.688  0.573 |  |  |
| *Religion:*  Christian  Muslim  Others | 12 (5.5)  39 (6.9)  0 (0.0) | Ref  1.3 (0.7 – 2.5)  (empty) | 0.466  (empty) |  |  |
| *Age at sexual debut:*  *10 – 14 years*  *15 – 17 years*  *18 – 24 years* | 9 (11.0)  22 (7.8)  18 (4.5) | Ref  0.7 (0.3 – 1.6)  0.5 (0.2 – 0.9) | 0.367  0.023 | Ref  0.5 (0.2 – 1.2)  0.4 (0.1 – 1.2) | 0.126  0.099 |
| *Marital status:*  Single /Dating/Cohabiting  Engaged/Married | 7 (5.5)  44 (6.6) | Ref  1.2 (0.5 – 2.8) | 0.634 |  |  |
| *Primary caregiver:*  None  Male partner  Parents  Relatives/Neighbors | 0 (0.0)  40 (6.4)  9 (6.3)  2 (14.3) | (empty)  0.4 (0.1 – 1.9)  0.4 (0.1 – 2.1)  Ref | (empty)  0.253  0.278 |  |  |
| *Age group of male partners:*  <25 years  25-29 years  30+ years | 9 (3.8)  26 (7.2)  12 (6.8) | Ref  1.9 (0.9 – 4.2)  1.8 (0.7 – 4.4) | 0.102  0.195 |  |  |
| *Employed or in business:*  No  Yes | 45 (6.3)  6 (8.1) | Ref  1.3 (0.5 – 3.2) | 0.547 |  |  |
| *Planned pregnancy:*  No  Yes | 13 (6.0)  36 (6.4) | Ref  1.1(0.6 – 2.1) | 0.844 |  |  |
| *Parity:*  Nulliparous  1  2  3+ | 25 (5.6)  11 (6.9)  12 (11.8)  3 (3.8) | Ref  1.3 (0.6 – 2.6)  2.3 (1.1 – 4.7)  0.7 (0.2 – 2.3) | 0.552  0.028  0.517 | Ref  0.9 (0.4 – 1.9)  1.3 (0.6 – 2.9)  0.2 (0.0 – 0.8) | 0.739  0.520  0.026 |
| *Age group at first pregnancy:*  14 – 17 years  18 – 19 years  20 – 24 years | 17 (7.3)  15 (5.6)  19 (6.0) | Ref  0.8 (0.4 – 1.5)  0.8 (0.4 – 1.6) | 0.444  0.543 |  |  |
| *Gestation at first ANC:*  0-13 weeks  14-19 weeks  20-24 weeks | 8 (8.3)  5 (3.6)  38 (6.9) | Ref  0.4 (0.1 – 1.3)  0.8 (0.4 – 1.8) | 0.126  0.607 |  |  |
| *Received any ANC information since becoming pregnant:*  No  Yes | 7 (2.2)  44 (9.3) | Ref  4.6 (2.0 – 10.2) | <0.001 | Ref  3.3 (1.4 – 8.2) | 0.008 |
| *Knowledge on when to start ANC clinic:*  On missing periods  1^st^ trimester  2^nd^ trimester  3^rd^ trimester  Don’t know | 10 (17.9)  24 (7.3)  15 (5.0)  0 (0.0)  2 (2.6) | Ref  0.4 (0.2 – 0.8)  0.2 (0.1 – 0.6)  (empty)  0.1 (0.0 – 0.6) | 0.013  0.001  (empty)  0.008 | Ref  0.3 (0.1 – 0.7)  0.3 (0.1 – 0.8)  (empty)  0.2 (0.0 – 0.9) | 0.004  0.021  (empty)  0.034 |
| *Knowledge on how many ANC visits is recommended:*  *None*  *Between 1-2*  *Between 3-5*  *At least 4*  *As many as possible* | 1 (4.4)  3 (8.6)  9 (4.7)  23 (9.4)  15 (5.1) | Ref  2.1 (0.2 – 21.1)  1.1 (0.1 – 8.9)  2.3 (0.3 – 17.8)  1.2 (0.1 – 9.3) | 0.542  0.942  0.428  0.876 |  |  |
| *Time to facility (mins):*  0-30  31-60  61+ | 15 (6.5)  17 (5.5)  19 (7.5) | Ref  0.8 (0.4 – 1.7)  1.2 (0.6 – 2.3) | 0.618  0.673 |  |  |

*ANC = Antenatal Care; Adjusted analyses include age, education, religion, age at sex debut, marital status, planned pregnancy, parity, and time to facility
